# A rapid review of barriers and facilitators to cancer screening uptake (breast, cervical and bowel) in underserved populations

**DOI:** 10.1101/2022.08.11.22278362

**Authors:** Chukwudi Okolie, Amy Hookway, Alesha Wale, Jordan Everitt, Hannah Shaw, Ruth Lewis, Alison Cooper, Adrian Edwards

## Abstract

In the United Kingdom (UK), the National Health Service (NHS) provides population-based screening programmes for breast, bowel, and cervical cancer. These programmes were temporarily paused in March 2020, due to the COVID-19 pandemic, resulting in large numbers of the eligible population having their invitations delayed. This disruption may have had a disproportionate impact on underserved populations for whom there was a lower uptake prior to the pandemic. Some people may also be less willing to attend screening after the pandemic. Interventions and campaigns designed to encourage people to take part in cancer screening may need to be adapted after the pandemic, in particular those targeting underserved populations.

This rapid review aimed to identify the barriers and facilitators to breast, bowel, and cervical screening uptake in underserved populations (e.g. clinically vulnerable, shielding, multi-morbidities, ethnic minorities, social deprivation, gender, age) during and since the onset of the pandemic, using evidence from the UK and other countries with similar cancer screening programmes (such as Australia and Netherlands), and to compare with the pre-pandemic literature. The pre-pandemic literature was identified using a supplementary scoping search for published systematic reviews.

Three primary studies (two published and one ongoing trial) conducted during the pandemic were identified. Five systematic reviews of pre-pandemic evidence were also included. Two qualitative studies conducted during the pandemic were appraised as high quality but both included sample populations with limited representation.

No primary studies specifically exploring the impact of the pandemic on barriers and facilitators to screening uptake among underserved groups were identified. The findings did not show marked differences in the barriers and facilitators for screening uptake before and during the COVID-19 pandemic in underserved populations. However, it is unclear whether this is because these genuinely remain unchanged or reflects the lack of available evidence. The findings may only be transferable to the population groups studied.

## 1. BACKGROUND

This Rapid Review is being conducted as part of the Wales COVID-19 Evidence Centre Work Programme. A number of research question on cancer screening participation were submitted during the Prioritisation process by multiple stakeholders. A preliminary meeting between interested parties was used to clarify what would be the most useful focus to influence policy. It was decided that this review would address the research question ‘Have the barriers and facilitators to cancer screening uptake (breast, cervical and bowel) in underserved populations (e.g. clinically vulnerable, shielding, multi-morbidities, ethnic minorities, social deprivation, gender, age), changed due to the COVID-19 pandemic?’

### 1.1 Purpose of this review

Cancer is a leading cause of death worldwide, accounting for approximately 10 million deaths in 2020 (World Health Organization, 2022). Population cancer screening services are used to reduce cancer mortality by offering asymptomatic people screening to detect precancerous lesions or cancers at an early stage. In the United Kingdom (UK), the National Health Service (NHS) provides population-based screening programmes for breast, bowel, and cervical cancer (NHS, 2021). Currently in Wales, breast screening is offered every three years to women aged 50 to 70 years; bowel cancer screening is offered every two years to people aged 58 to 74 years, and cervical screening is offered every five years to women aged 25 to 64 years (Public Health Wales, 2022). Due to the COVID-19 pandemic, these programmes were temporarily paused in March 2020, leaving large numbers of participants without access to these routine services.

Prior research has reported low uptake of cancer screening programmes by individuals from socioeconomically deprived communities and among ethnic minority and underserved populations (Nardi et al., 2016, Ni et al., 2020, Szczepura et al., 2008). Higher rates of non-attendance in cancer screening programmes within these population groups are therefore likely to result in more cancer diagnoses and cancer related deaths within these already disadvantaged communities (Wearn and Shepherd, 2022). Underserved population groups have also been disproportionately impacted by the COVID-19 pandemic, which may lead to even wider inequalities in cancer screening uptake within these groups.

There is increasing concern regarding the effect of the coronavirus pandemic on cancer mortality, particularly among underserved populations. Given the perceived need to address health inequalities in these groups, understanding the barriers and facilitators to participation in screening programmes is important. The purpose of this rapid review is to identify the barriers and facilitators to breast, bowel, and cervical screening uptake in underserved populations during and since the onset of the pandemic, using evidence from the UK and other countries with similar cancer screening programmes (such as Australia and Netherlands), and to compare with the pre-pandemic literature.

## 2. RESULTS

### 2.1 Overview of the Evidence Base

#### 2.1.1 Pandemic-related evidence

Three primary studies were identified for inclusion in this rapid review. These included two published qualitative studies (Christie-de Jong et al., 2022, Keane et al., 2022) and one ongoing randomised controlled trial (RCT) (Wilding, 2022).

The two published qualitative studies targeted specific underserved or deprived population groups. One study (Christie-de Jong et al., 2022), which was conducted in the UK, focused on Muslim women and participation in breast, bowel and cervical cancer screening. The second study (Keane et al., 2022), which was conducted in the Republic of Ireland, focused on Irish Traveller women and participation in breast cancer screening.

Both published studies explored factors affecting the uptake of cancer screening programmes. One study investigated barriers to screening uptake while the other investigated both barriers and facilitators to uptake. Despite being conducted during the COVID-19 pandemic, neither study focused specifically on the impact of the pandemic on perceptions towards cancer screening among underserved groups.

The methodological quality of the two published primary studies were assessed using the 10-item Critical Appraisal Skills Programme (CASP) tool for qualitative research (Critical Appraisal Skills Programme, 2018). Both studies were deemed to be of high quality as they met the majority of the CASP tool criteria (see Appendix 3). However, it is worth noting the limitations of both studies including: study participants in Keane et al (2022) were working as Traveller Primary Health Care workers at time of interview, and participants in Christie-de Jong et al (2022) were self-selected, educated and English-speaking, with possibly fairly positive attitudes to screening already.

A summary of the two qualitative studies is provided in Tables 1, and a narrative summary of their findings are presented in Section 2.2.

**Table 1.**
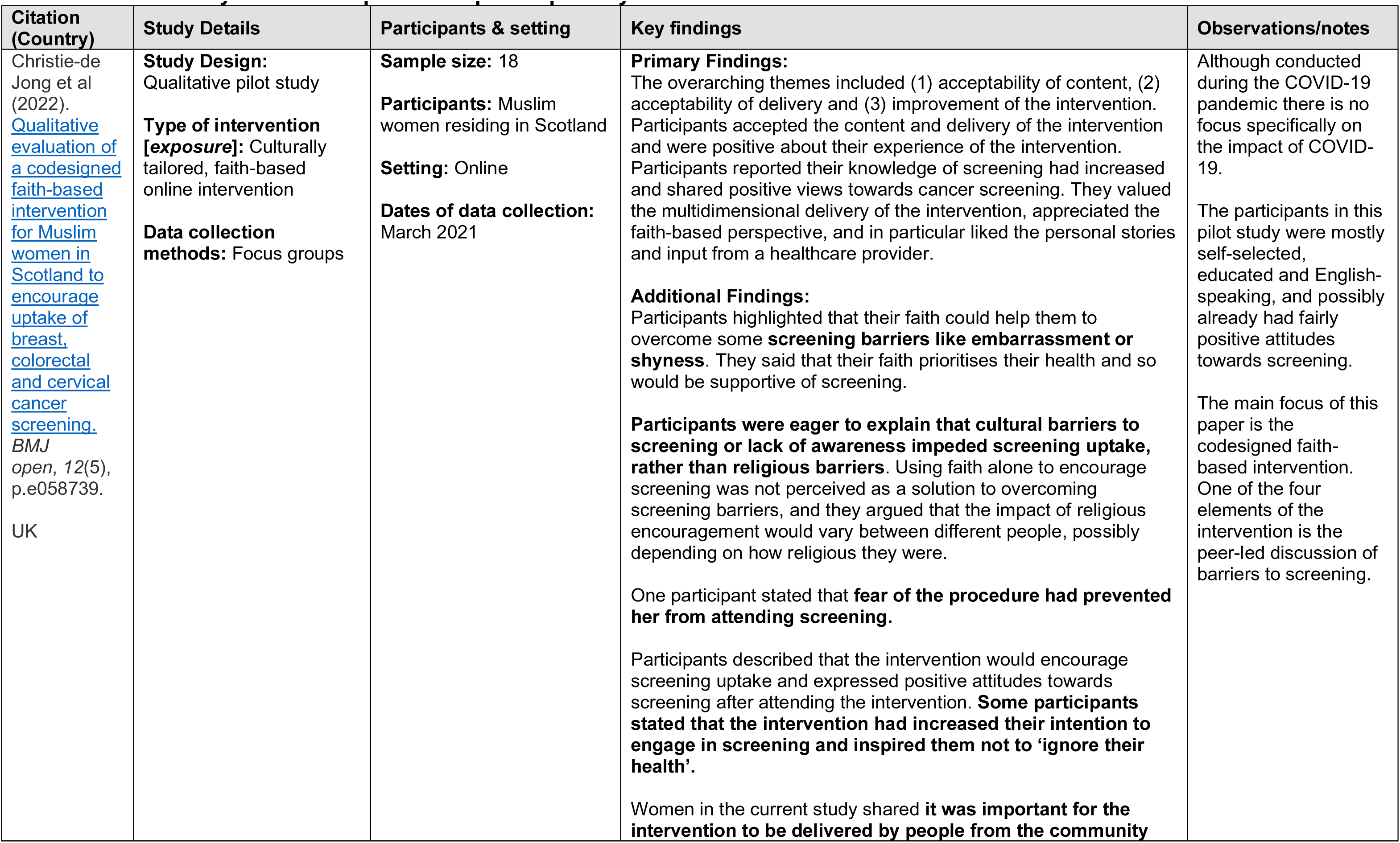
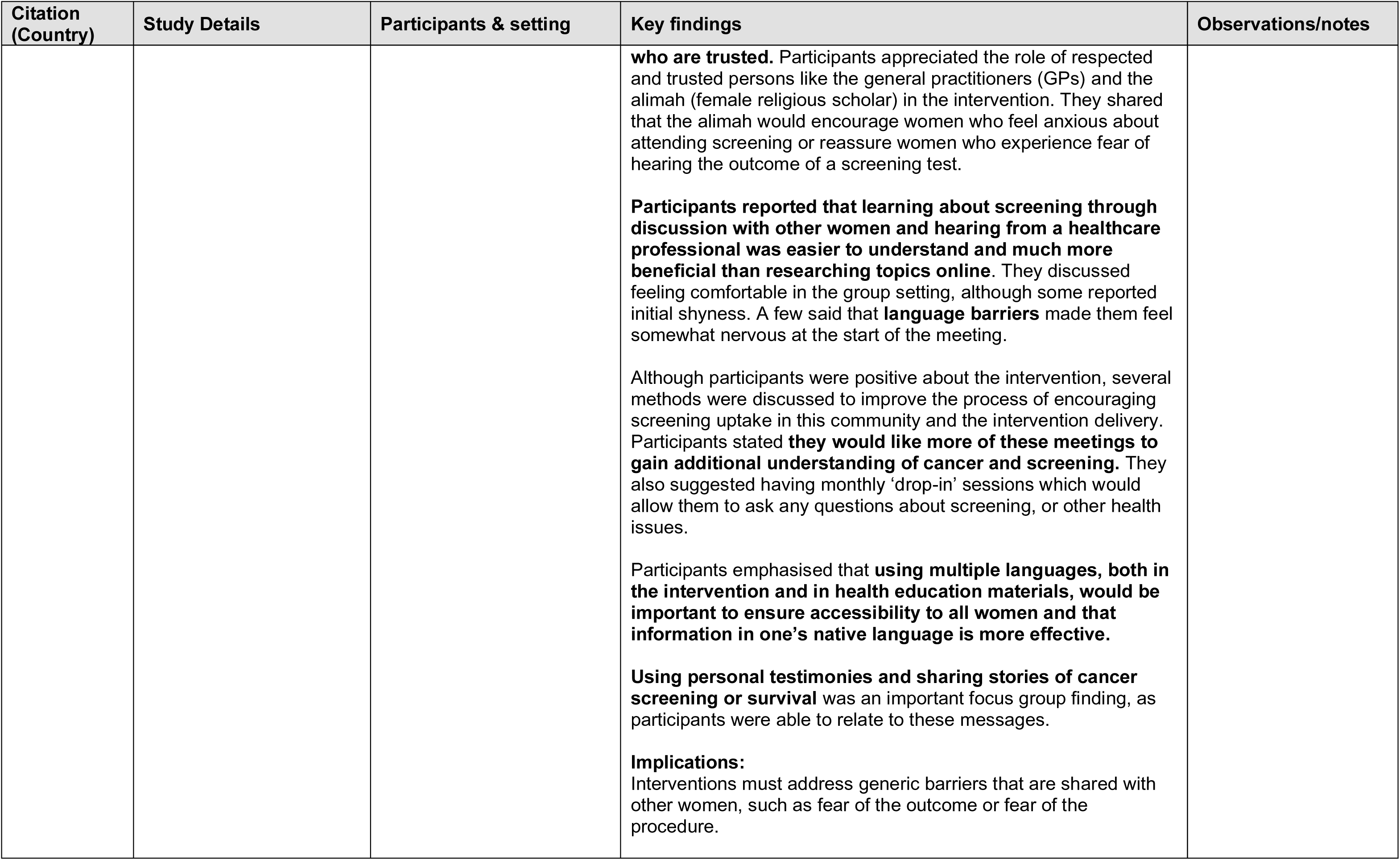
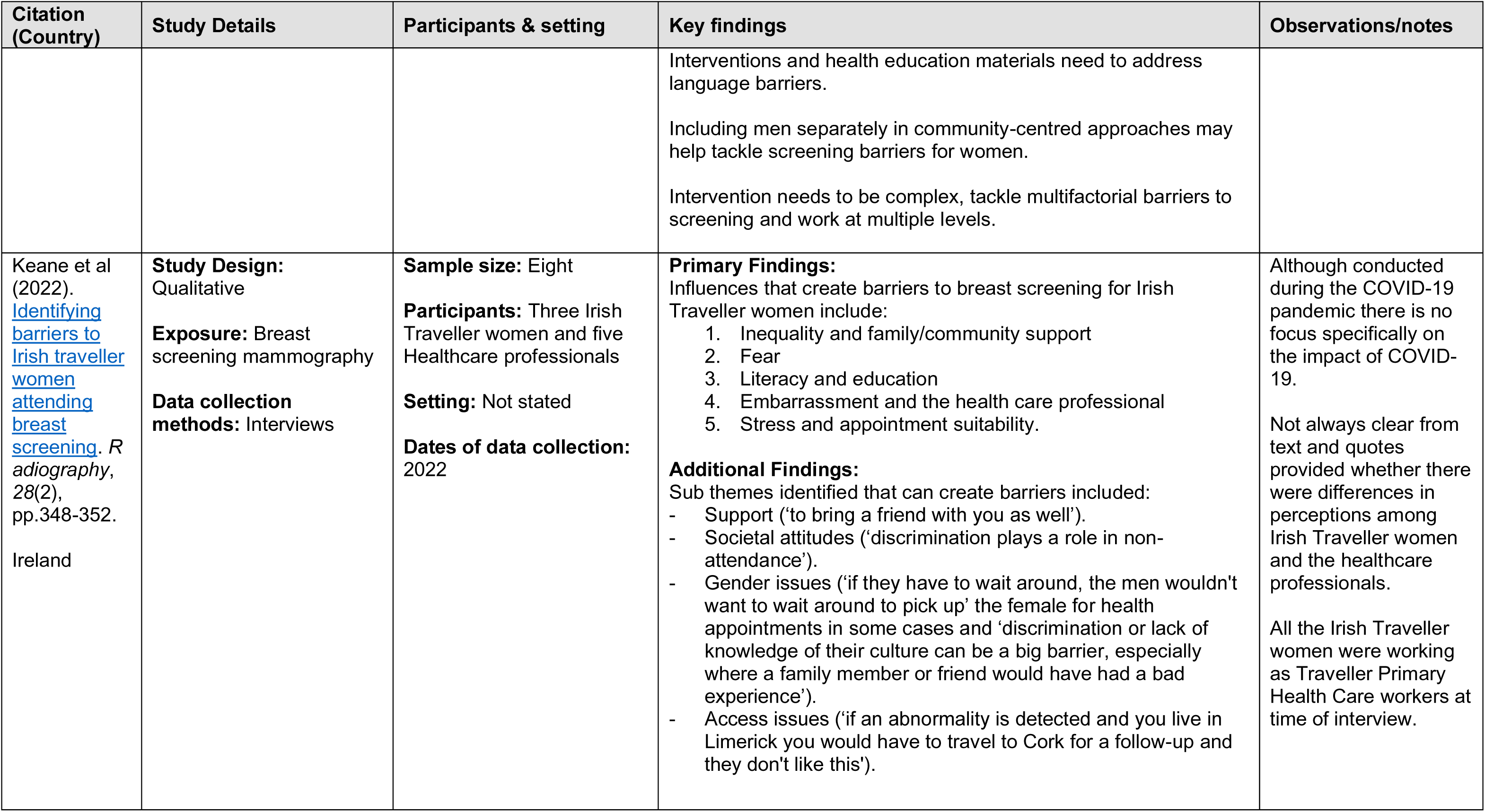
Summary of included pandemic-period primary studies

The ongoing RCT focuses on cervical screening uptake among both deprived and non-deprived groups in the UK. A summary of the trial is provided in Table 2. No results have yet been reported for this study.

**Table 2.**
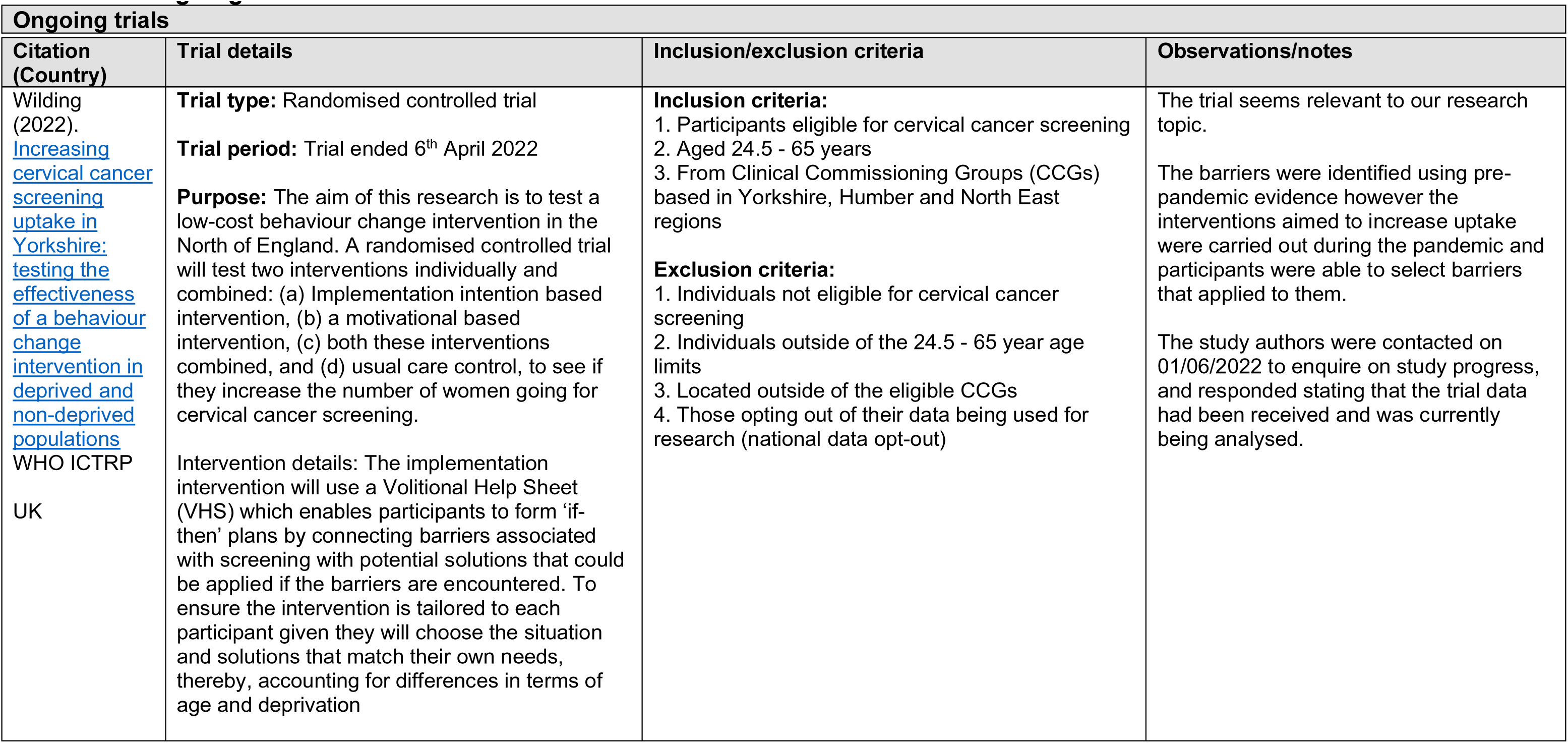
Ongoing trials

#### 2.1.2 Pre-pandemic evidence

This rapid review aimed to identify studies conducted during and since the onset of the pandemic. The pre-pandemic literature used here for comparison was identified using a supplementary scoping search for published systematic reviews (see Section 5). Five systematic reviews (Baird et al., 2021, Travis et al., 2020, Wearn and Shepherd, 2022, Wessex Voices, 2021, Young et al., 2018) reporting pre-pandemic evidence on barriers and facilitators to breast, bowel, and cervical screening uptake in various underserved populations were identified. These reviews sought the barriers and facilitators among Black, Asian and minority ethnic (BAME) groups, low-uptake sociodemographic groups, people with mental illness or learning disabilities, people with physical disabilities, Lesbian, Gay, Bisexual and Trans (LGBT) people and underserved women. Two reviews focused on breast screening, one on bowel screening, one on cervical screening, and one on breast, bowel, and cervical screening. Primary studies included in the systematic reviews were mostly qualitative and conducted in the UK.

The methodological quality of the five pre-pandemic systematic reviews were assessed using the AMSTAR 2 critical appraisal tool (Shea et al., 2017). Four reviews were rated as being of critically low quality, while one review was rated as low quality.

A summary of included evidence is provided in Table 3. Narrative summaries of the evidence are presented below.

**Table 3.** Summary of included pre-pandemic secondary studies

| Citation | Review details | Included studies | Quality | Findings and observations/notes |
| --- | --- | --- | --- | --- |
| Baird et al (2021). What can be done to encourage women from Black, Asian and minority ethnic backgrounds to | **Review period:** Up to December 2017<br>**Review purpose:** To identify the barriers to UK Black, Asian, and Minority Ethnic women attending breast screening, and | **Number of included studies:** 8<br>**Key characteristics:**<br>All included qualitative studies were UK-based and concerned Black, Asian, and Minority Ethnic women | Critically low | **Knowledge-related factors:**<br>• Lack of knowledge on: what is breast cancer, how to identify it, what is the screening programme, who is at risk and the treatments available.<br>**Cultural-related factors:**<br>• Cultural values |
| attend breast screening? A qualitative synthesis of barriers and facilitators. Public Health: DOI: 10.1016/j.puhe.2020.10.013 | solutions to the public health problem of ethnic variation within screening attendance.<br>**Included study designs:**<br>Qualitative<br>**Included outcome measures:**<br>Barriers and facilitators to breast cancer screening | participants. The majority of studies were based in London. |  | Religious beliefs (associations between faith and decreased appreciation of preventative medicine, with women describing the development of breast cancer as 'up to God')<br>Influence of family and friends<br>Stigma associated with cancer<br>Fear of mastectomy and its marital consequences<br>Deficient support from family and community<br>Gender of healthcare professionals (male radiographer made some women reluctant to partake)<br>Previous negative experiences with healthcare professionals (exacerbated in situations where women felt disrespected)<br>**Access-related factors:**<br>Distance<br>Inconvenience<br>Cost<br>**Facilitators:**<br>Educational facilitators – improve knowledge<br>Incorporation of religious leaders into educational interventions<br>Offering language support<br>Forming interpersonal relationships between healthcare workers and BAME<br>Female radiographer (unanimously preferred across all ethnicities)<br>Encouragement from healthcare professionals<br>Supportive families and communities<br>Transport provision<br>Positive health messages from health services<br>Information leaflets in different languages |
| Travis et al (2020). Barriers to flexible sigmoidoscopy | **Review period:** From database inception to January 2020<br>**Review purpose:** To synthesise qualitative evidence related to | **Number of included studies:** 10<br>**Key characteristics:** | Low | **Barriers:**<br>Procedural anxieties - anxieties regarding test invasiveness (Women and UK Asian communities)<br>Shame and embarrassment (UK Asian groups) |
| colorectal cancer screening in low uptake socio-demographic groups: A systematic review.<br>Psychooncology.<br>doi:<br>10.1002/pon.5443<br>. | barriers and facilitators of flexible sigmoidoscopy screening (FSS) intention and uptake, particularly within low socio-demographic uptake groups.<br>**Included study designs:**<br>Qualitative<br>**Included outcome measures:**<br>Barriers and facilitators of FSS intention and uptake | Of the ten included studies, six were from the UK, three from USA, and one study from Canada.<br>Qualitative data collection methods included focus groups (n = 3), telephone semi-structured interviews (n = 2), and face-to-face semi-structured interviews (n = 5). The method of analysis carried out by many of the included studies was thematic framework analysis. |  | • Masculinity-associated procrastination/machismo. FSS considered as a threat to masculinity (Afro-Caribbean men)<br>• Reduced perceived personal susceptibility to CRC (Residents in deprived area)<br>• Medical fear of doctors, hospitals, and tests in general<br>• Anxiety surrounding test results<br>• Avoidance due to underlying fatalism<br>• Belief that treatment alone caused cancer to advance (Pakistani women)<br>• Anticipation of fear and anxiety (Gujarati Indian men)<br>• Perceived susceptibility to CRC (Gujarati Indian men)<br>• Lack of awareness of prevalence (Gujarati Indian men) and lack of knowledge about CRC (Pakistani women)<br>• Procedural pain and discomfort anticipated and experienced from FSS<br>• Women's experience of painful mammograms heightened nervousness to attend the FSS test<br>• Peer pressure, a lack of family support or encouragement were found to both promote and inhibit screening intention and uptake<br>• Religious and cultural-influenced health beliefs<br>• Competing priorities<br>**Facilitators:**<br>• Peer pressure, a lack of family support or encouragement were found to both promote and inhibit screening intention and uptake<br>• Doctor/physician screening recommendation, in which personalised invitations from medical professionals promoted screening<br>• Sense of responsibility to use public funding and resources<br>• Presence of a professional throughout the procedure |
| Wearn and Shepherd (2022). Determinants of | **Review period:** Searches were conducted in June 2018 and repeated in January 2021 | **Number of included studies:** 24<br>**Key characteristics:** | Critically low | **Barriers:**<br>• Embarrassment - procedure is physically and emotionally invasive |
| routine cervical screening participation in underserved women: a qualitative systematic review.<br>Psychology & Health. DOI: 10.1080/08870446.2022.2050230 | **Review purpose:** To synthesise qualitative literature which explore determinants of cervical screening participation within the context of population wide call-recall screening programmes. Specifically, this review aims to synthesise the views of ethnic minority women and women living in socioeconomically deprived communities, identifying key commonalities across these underserved groups<br>**Included study designs:**<br>Qualitative<br>**Included outcome measures:**<br>Determinants of cervical screening participation | Of the 24 included studies, 10 were from the UK, seven from Australia, three from the Netherlands, two from Norway, and one each from Finland and Sweden.<br>The majority of studies (n = 16) were focused upon migrant women. Two further studies grouped women as a minority based upon their religious beliefs, three studies classified participants as an ethnic minority if they self-defined as an ethnic group other than the country's majority population and one study described participants as an ethnic minority in relation to their language group. Two studies classified participants as low socioeconomic status as they lived within areas of high deprivation |  | • Religious beliefs - aspect of shame associated with 'exposing' oneself, belief that God would keep them safe from cancer, perceived as God's will<br>• Fear of the test and potential outcome<br>• Risk beliefs (perceived low risk)<br>• Religious beliefs<br>• Prioritising competing demands<br>• Perceived stigma – judgements, perceived as sexually active, perceived as a cause of promiscuity (religious concerns)<br>• Lack of knowledge of purpose of screening<br>• Peer and family influence – negative stories, partners views<br>• Barriers to communication<br>• Unfamiliarity with screening, differences to home country, not mandatory therefore maybe seemed unimportant<br>• Negative past experiences of healthcare and screening<br>• Sex of practitioner, preference for female<br>• Interpersonal skills of practitioner<br>• Continuity of care, see a practitioner they had built rapport with<br>• Medical mistrust<br>• Practitioner endorsement (by a known practitioner)<br>• Service accessibility, time, travel, childcare<br>• Cultural differences, preventive health perceived as searching for disease which may trigger ill-health, female circumcision and fear of judgement<br>**Facilitators:**<br>• Receipt of invitation letter<br>• Practitioner endorsement<br>• Free-of-charge test<br>• Media promotion, e.g., human interest stories, raising awareness and encouragement |
|  |  |  |  | • Strong social networks and close family members<br>• Perceived severity of cancer<br>• Relief of receiving positive outcome<br>• Friendly and approachable healthcare staff |
| Wessex Voices (2021). A systematic review of people's experiences of breast screening - A rich and diverse picture. | **Review period:** 2000 to 2021<br>**Review purpose:** To understand the views of people eligible for breast screening and identify gaps in knowledge and areas where further insight is needed<br>**Included study designs:** Not stated<br>**Included outcome measures:** Experiences of screening, barriers and facilitators/solutions | **Number of included studies:** 105<br>**Key characteristics:** Not stated | Critically low | **Ethnic minorities:**<br>**Barriers:**<br>• Difficulties accessing information and the breast screening programme<br>• Culturally appropriated information<br>• Screening in native country viewed as superior (more regular)<br>• Information doesn't address specific concerns of ethnic minority groups<br>• Past negative experience<br>• Language difficulties<br>• Lack of awareness of system in UK<br>• Lack of knowledge, who's at risk, how do you get it<br>• Lack of access to information<br>• Use of incomprehensible terminology on NHS materials<br>• Not comfortable exposing breasts to a stranger<br>• Religious beliefs, e.g., cancer was a retribution for past actions and that illness was a means of forgiveness - hence not seeking treatment. Screening non-diagnostic, rules allowing exposure of sensitive areas do not apply<br>• Embarrassment<br>• Don't want a male practitioner<br>• Lack of support from family and community<br>• Fear of diagnosis<br>• GPs able to exclude people with mental illness from being invited to screening<br>• Locations, e.g., supermarket car parks, can be embarrassing<br>**Facilitators:** |
|  |  |  |  | Knowledge around breast screening<br>Translated screening materials & letter<br>**For people with mental illness or learning disabilities:**<br>**Barriers:**<br>Negative past experiences<br>Sensory overload<br>Fear and anxiety<br>Need to convince carers who may make decisions for patients<br>**Physical disabilities:**<br>**Barriers:**<br>Access, steps, stairs, small screening rooms, lifts, distance from parking location<br>Physical limitations using screening equipment<br>**Lesbian, Gay, Bisexual and Trans:**<br>**Barriers:**<br>No automated call-recall system because the system is still defined by binary genders – male or female<br>Overt gendering or ‘pinking’ of breast and gynae cancers can be a barrier for LGBT people seeking information |
| Young et al (2017). Factors influencing the decision to attend screening for cancer in the UK: a meta-ethnography of qualitative research. Journal | **Review period:** Database inception to October 2016<br>**Review purpose:** To better understand experiences of being invited to cancer screening and associated decision-making<br>**Included study designs:** Qualitative | **Number of included studies:** 34<br>**Key characteristics:**<br>All included studies were from the UK. Twenty-one papers had cancer screening uptake as the main focus of the reports. The primary focus of other reports included wider knowledge and attitudes to cancer and prevention, responses to | Critically low | **Findings in ethnic minority and/or socioeconomically diverse population groups:**<br>**Breast cancer screening barriers:**<br>Time, travel, competing time demands<br>Language barrier<br>Cultural beliefs<br>Lack of confidence in screening and outcome<br>Relationship with health professionals |
| of Public Health.<br>doi:<br>10.1093/pubmed/fdx026 | **Included outcome measures:**<br>Factors influencing cancer screening attendance | information about screening, experiences of screening test results and risk management options which included screening. Cervical, breast and colorectal cancer accounted for 29 of the 34 studies. Two related to prostate cancer, two to ovarian and one to lung cancer. |  | Religious beliefs<br>Service provision issues<br>The test itself<br>Apathy<br>Low-risk perceptions<br>**Facilitators:**<br>Invitation letter<br>Belief that screening is effective and beneficial<br>**Cervical cancer screening barriers:**<br>Lack of knowledge (about cervical cancer, its risk factors, causes and screening)<br>Lack of awareness of rights to healthcare in Britain<br>Language difficulties<br>Fear of the test or pain<br>Fear of cancer<br>Embarrassment<br>Shame<br>Negative past experiences<br>Male practitioners<br>Practical difficulties, location/travel, time, childcare<br>Absence of symptoms / perceived risk<br>Perceived as an inconvenience<br>Lack of confidence in NHS health professionals<br>'Fractured living' in two countries (lack of acculturation)<br>Cultural beliefs e.g. cervical cancer associated with promiscuity, inflicted as a punishment from God, a disease of the West, nothing could be done to avoid cervical cancer<br>Racism (treated coldly because of race, being treated like a piece of meat, being too intimidated to ask questions)<br>**Colorectal cancer:**<br>Lack of knowledge / awareness |
|  |  |  |  | • Invasiveness of the test and the area of the body under investigation<br>• Psychosocial barriers to bowel preparation at home (enema)<br>• Fear of test results<br>• Lack of symptoms<br>• Biomedical view of healthcare system<br>• Language difficulty<br>• Threat to masculinity<br>• Knowing a close family member or friend who had died of cancer<br>• Feeling susceptible<br>• Stigma of cancer<br>• Embarrassment<br>• Belief that God would help them<br>• The word 'occult' having demonic connotations<br>• Attitudes of staff to religious beliefs e.g., female endoscopist necessary<br>**Barriers to faecal occult blood test completion:**<br>• Everyday pressures<br>• Faecal sample<br>• Misunderstanding of instructions<br>• Planning test completion<br>• Risks that collecting, storing, and posting samples of faeces posed to hygiene<br>• Preference to attend a health setting<br>• Risk perception<br>• Participants preferred not to be in possession of this information (screening results)<br>**Findings in people with mental illness (breast, cervical and bowel screening):**<br>**Barriers:** |
|  |  |  |  | • Not knowing what to expect or what to do; unsure of need for screening; difficult to process information<br>• Lack of understanding of mental illness in screening professionals; made to feel like a burden on health service; stigma of mental illness<br>• Screening environment aggravates mental health symptoms; staff can be rushed; staff can be rough; exclusion from GP registers<br>• Additional burden; mental health symptoms reduce motivation for self-care; past negative experience; embarrassment; traumatizing; fear of bad news; poor relationship with GP; diagnostic overshadowing<br>• Appointment booking; transport difficulties; difficulty remembering appointments; difficulty leaving the house due to mental health problems; taking time off<br>**Facilitators:**<br>• Wanting to be informed; understanding the benefits of screening; encouragement<br>• Staff being understanding; staff knowledge of mental illness<br>• Continuity of care<br>• Feeling health conscious; being anxious to avoid further health problems; physical symptoms (e.g. finding a lump); past positive experience; good relationship with GP; good relationship with practice nurse<br>• Familiar location; reminders |

### 2.2 Barriers and facilitators to cancer screening uptake

#### 2.2.1 Pandemic-related evidence

Keane et al. (2022) conducted a study to assess the perceptions of Irish Traveller women towards breast screening and the perceived barriers and enablers to attendance. Findings identified influences creating barriers to breast screening for this group including **inequality and family or community support, fear of a negative outcome, literacy and education, embarrassment and the health care professional, and stress and appointment suitability**.

Christie-de Jong et al. (2022) conducted a pilot qualitative study aimed at evaluating the acceptability of a co-designed faith-based intervention to encourage uptake of breast, colorectal and cervical cancer screening in Scottish Muslim women. While participants accepted the content and delivery of the intervention, they also highlighted barriers to cancer screening including **embarrassment/shyness, fear of the procedure/outcome, lack of awareness, social stigma surrounding colorectal, breast, and cervical cancers, and cultural and language barriers**. Participants were also eager to explain that cultural barriers to screening or lack of awareness impeded screening uptake, rather than religious barriers.

Facilitators to breast, colorectal and cervical screening uptake included the **use of faith-based interventions delivered by people from the community who are trusted, the use of multiple languages both in interventions and in health education materials and the sharing of personal testimonies and stories of cancer screening or survival during focus group meetings**.

#### 2.2.2 Pre-pandemic evidence

Baird et al. (2021) conducted a systematic review to identify barriers and facilitators to breast screening attendance in UK BAME women. The main barriers identified included knowledge-related factors such as, **lack of knowledge surrounding breast cancer and screening, and language barriers**; Cultural-related factors such as, **stigma associated with cancer, fear of mastectomy and its marital consequences, deficient support from family and community, gender of healthcare professionals, faith and under-appreciation of preventative medicine, and cultural beliefs related to modesty**; and Access-related factors such as, **logistical (distance, inconvenience and cost) and emotional barriers.**

**Facilitators included educational facilitators such as,** incorporation of religious leaders into educational interventions, improving knowledge, offering language support, forming interpersonal relationships between healthcare workers and BAME women, encouragement from healthcare professionals, and supportive family members and communities**; and Logistic facilitators such as,** transport provision, positive health messages from health services, and use of information leaflets in different languages.

Travis et al. (2020) conducted a systematic review on barriers and facilitators of flexible sigmoidoscopy screening (FSS) intention and uptake within low socio-demographic uptake groups (women, lower socioeconomic status (SES), and Asian minority ethnicity).

**Barriers included** procedural anxieties, ‘medical fear’ of doctors, hospitals, and tests, shame and embarrassment, masculinity-associated procrastination/machismo, anxiety surrounding test results, perceived susceptibility to colorectal cancer (CRC), lack of knowledge and awareness about CRC, and religious and cultural-influenced health beliefs.

Facilitators included **sense of responsibility to use public funding and resources, presence of a professional throughout the procedure, family support and encouragement, doctor/physician screening recommendation, and personalised invitations from medical professionals**.

Wearn and Shepherd (2022) conducted a qualitative systematic review on the determinants of routine cervical screening participation in underserved women. Barriers to participation included **embarrassment, fear of the test and potential outcome, risk beliefs, religious beliefs, prioritising competing demands, perceived stigma, lack of knowledge, peer and family influence, communication barriers, unfamiliarity with screening, negative past experiences of healthcare and screening, gender of practitioner, interpersonal skills of practitioner, medical mistrust, service accessibility, and cultural differences.**

**Facilitators to participation included** perceived severity of cancer, anticipated relief of receiving a positive outcome, strong social networks, close family members, friendly and approachable healthcare staff, practitioner endorsement, service accessibility, and media influence on raising awareness.

Wessex Voices (2021) conducted a systematic review on people’s experiences of breast screening. The main barriers to breast screening uptake in ethnic minorities identified were **lack of knowledge, language difficulties, use of incomprehensible terminology on NHS materials, lack of confidence and expectation anxiety, fear, stigma associated with cancer, embarrassment, religious beliefs**.

Barriers for people with mental illness or learning disabilities included **negative past experiences, and fear and anxiety**. Barriers for people with physical disabilities included **lack of disability access and inaccessibility of equipment.** Barriers for LGBT people included **discrimination, misgendering, and lack of provider awareness.**

Young et al. (2017) conducted a review aimed at understanding the factors influencing the decision to attend screening. Barriers to breast cancer-screening uptake in ethnic minority and/or socioeconomically diverse groups included **language barriers, cultural beliefs, apathy, low risk perception, competing time demands, and religious beliefs.**

Barriers to cervical cancer screening included **lack of knowledge/awareness, language difficulties, fear of the test, fear of cancer, embarrassment and shame, negative past experiences, male practitioners, cultural beliefs, and racism**.

Barriers to colorectal cancer screening included **lack of knowledge/awareness, invasiveness of the test, fear, language difficulties, threat to masculinity, stigma, embarrassment, and religious beliefs.**

Barriers to breast, cervical, and bowel screening uptake in people with mental illness included **stigma of mental illness, transport difficulties, fear of bad news, and not knowing what to expect**.

Facilitators to breast, bowel, and cervical screening uptake by people with mental illness included **staff being understanding, good relationship with practice staff, familiar locations, understanding the benefits of screening, and positive past experience.**

#### 2.2.3 Bottom line results for barriers and facilitators to breast, bowel, and colorectal cancer screening uptake

##### 2.2.3.1 Pandemic-period

Evidence around the barriers and facilitators to cancer screening uptake in underserved populations during the pandemic is limited. However, from the limited evidence available, barriers among Scottish Muslim women and Irish Traveller women include **lack of support from family and community, fear of the procedure/negative outcome, embarrassment, gender of healthcare professional, lack of education, lack of awareness, appointment suitably, social stigma surrounding colorectal, breast, and cervical cancers, and cultural and language barriers.** Certain barriers may be specific to each underserved population, with others common across the underserved groups. Conversely, facilitators to uptake appear to include **trust, offering language support, and use of personal testimonies**.

##### 2.2.3.2 Pre-pandemic period

Evidence from the pre-pandemic period appears to be much more abundant and indicated that barriers to cancer screening uptake in underserved populations (such as people with mental illness, ethnic minorities, underserved women, low-uptake sociodemographic groups) included a **lack of knowledge and awareness; language and communication barriers; perceived stigma associated with cancer; lack of support from family and community; religious and cultural beliefs; logistical barriers; fear and anxiety; shame and embarrassment; gender of practitioner; medical mistrust; machismo/threat to masculinity; discrimination and racism; misgendering; and negative past experience**.

Facilitators to cancer screening uptake identified from the literature included **positive past experience, encouragement from healthcare professionals, supportive family members and communities, transport provision, offering language support, friendly and approachable healthcare staff, practitioner endorsement, and educational support.**

## 3. DISCUSSION

### 3.1 Summary of the findings

A comparison of evidence on barriers and facilitators to breast, bowel and cervical cancer screening uptake in underserved populations, identified before and during the COVID-19 pandemic does not appear to show any marked differences. However, variation in the population groups as well as the type of screening undertaken, could impact on the generalisability of these findings.

Our rapid review has identified several barriers and facilitators to breast, bowel and cervical screening uptake. It is possible some may be unique to the individual population groups examined, but some, such as ‘fear’ (of procedure or outcome) appear to be a common barrier across most underserved populations. The barriers and facilitators highlighted appear similar across the different screening programmes investigated. In light of this, the findings may only be generalisable to the populations groups they were identified in, and possibly the type of cancer screening.

It is important to note that our rapid review focused on research undertaken in a few select countries (Australia, Netherlands, Republic of Ireland and UK)as these are comparable with UK cancer screening programmes. We however identified one study conducted in The Republic of Ireland which has similar screening criteria to the UK. It is likely we have missed several factors influencing uptake of cancer screening, and those identified are in no way an exhaustive list.

The ongoing UK trial identified in our searches reports an intention to investigate the barriers associated with cervical cancer screening uptake in order to test the effectiveness of a behaviour change intervention. Although we are unsure when the findings from this trial will be published (*study authors stated in recent correspondence that trial data is currently being analysed*), this trial may be relevant to this review topic and could likely contribute to the current body of evidence.

### 3.2 Limitations of the available evidence

There appears to be a paucity of UK (and countries with similar screening programmes) research evidence on cancer screening uptake in underserved groups conducted during the COVID-19 pandemic period. We identified some UK primary studies published during the pandemic, however, participant data were collected prior to the pandemic and therefore did not meet the inclusion criteria for this rapid review. In addition, we identified numerous studies from other countries, particularly USA, where data collection occurred during 2020 and 2021. These were not eligible for inclusion in this rapid review owing to differences in healthcare provision and screening criteria.

Various underserved population groups were identified in this rapid review, and it is highly possible barriers and facilitators may be unique to the different population groups. This is likely to impact on the generalisability of findings between population groups.

Although conducted during the COVID-19 pandemic, none of the primary studies identified specifically explored the impact of the pandemic on barriers and facilitators to screening uptake among underserved groups.

Likewise, some studies identified focused on the impact of the COVID-19 pandemic on people’s participation in cancer screening programmes. However, these were not targeted at underserved populations and therefore did not meet our inclusion criteria.

Although our findings show that barriers and facilitators to screening appear to remain unchanged before and during the pandemic, it is unclear whether these findings were due to the paucity of UK research evidence identified or the limited number of studies targeted at underserved populations.

### 3.3 Implications for policy and practice

It is likely that the evidence base around the impact of the pandemic on cancer screening uptake will continue to evolve as more studies publish results of their research. However, the findings from this review can serve as a useful baseline for future research to build on. They also serve as an evidence-based foundation for work to increase uptake and reduce inequity in uptake within the Screening Division of Public Health Wales.

Interventions designed to encourage uptake of cancer screening in underserved populations need to take into consideration the varied and complex nature of these groups. The evidence also indicates that as the barriers and facilitators to cancer screening are potentially different among each of the underserved groups, interventions may need to be tailored to individual population groups.

In light of the paucity of evidence identified, further well-designed higher quality research from the UK and similar countries is needed to better understand the factors affecting uptake of breast, bowel and cervical screening in underserved populations.

### 3.4 Strengths and limitations of this Rapid Review

The studies included in this rapid review were identified through an extensive search of electronic databases, trial registries, grey literature, as well as consultation of content experts in the field. Full text screening was conducted by one independent reviewer and any decision to include was checked for accuracy by a second reviewer. The data extraction was performed by one reviewer and two independent reviewers carried out consistency checking.

Rigorous review methods were used in this rapid review. Despite making every effort to capture all relevant publications and reduce the risk of bias, it is possible that additional eligible publications from may have been missed or we may have introduced some biases to this review.

Our decision to only include studies from countries where cancer screening programmes are closely comparable to UK (Australia, Netherlands, Republic of Ireland), means our findings are highly generalisable to the Welsh context. However, had we broadened our inclusion criteria to include other countries, it is likely we would have identified more barriers and facilitators. Whilst this may have increased our certainty of the evidence, it would have compromised our ability to generalise findings to the Welsh context.

Pre-pandemic evidence on barriers and facilitators to screening uptake were derived from secondary sources (systematic reviews) while evidence relating to the pandemic were derived from primary studies. This approach was chosen because our earlier Rapid Evidence Summary (RES) did not identify any relevant good quality pandemic-related secondary research. However, it should be noted that direct comparison of the evidence between systematic reviews and primary studies may not be appropriate as evidence from systematic reviews are normally ranked higher than that derived from primary studies.

The AMSTAR 2 tool was used to assess the quality of included pre-pandemic systematic reviews. This checklist, although validated, was designed specifically to assess the methodological quality of systematic reviews that include randomised or non-randomised studies of healthcare interventions. As such, it may not be the most suitable for appraising systematic reviews of qualitative studies.

## Data Availability

All data produced in the present study are available upon reasonable request to the authors

## 5. RAPID REVIEW METHODS

### 5.1 Eligibility criteria

We searched for primary sources to answer the review question “Have the barriers and facilitators to cancer screening uptake (breast, cervical and bowel) in underserved populations, changed due to the COVID-19 pandemic?”

The following eligibility criteria were used to identify studies for inclusion in the rapid review:

|  |  |  |
| --- | --- | --- |
| **Review question** | Have the barriers and facilitators to cancer screening uptake (breast, cervical and bowel) in underserved populations, changed due to the COVID-19 pandemic? |  |
|  | **Inclusion criteria** | **Exclusion criteria** |
| **Population** | Underserved populations (e.g. clinically vulnerable, shielding, multi-morbidities, ethnic minorities, social deprivation, gender, age) offered breast, bowel and cervical screening<br>General population if sub-group analyses includes an examination of underserved populations |  |
| **Exposure** | Breast, bowel and cervical screening programmes relevant to the UK context, during the COVID-19 pandemic | Other screening programmes<br>Screening programmes pre-pandemic |
| **Outcomes** | Barriers and facilitators to screening uptake | Screening uptake generally during the pandemic |
| **Settings** | Community and Hospital settings |  |
| **Studies** | Primary studies and grey literature. COVID literature (2020-22) |  |
| **Language of publication** | English |  |
| **Publication type** | Published and preprint, protocols |  |
| **Other factors**<br>*Any other key points to note* | **Only include studies conducted in the UK or other countries that have comparable cancer screening programmes to the UK (such as Australia and Netherlands)**<br>Limit search dates to 2020 onwards to capture COVID-19 relevance |  |

### 5.2 Literature search

COVID-19 specific and general repositories of evidence noted in our resource list were searched between 25^th^ and 26^th^ May 2022. An audit trail of the search process is provided within the resource list (Appendix). Searches were limited to English-language publications and limited to records published during the pandemic period (2020-2022). Search hits were screened for relevance by a single reviewer.

Search concepts and keywords around breast, bowel, and cervical cancer screening programmes, underserved populations, and barriers and facilitators were utilised. The searches combined free text words and descriptors when available. Resources searched during the rapid review are outlined in Appendix 1 and the search strategy used to search MEDLINE is available in Appendix 2.

A scoping search was also conducted to identify secondary sources relating to the pre-pandemic period, which are included in table 3. No date limits were applied, however, searches were limited to English-language publications and to secondary sources published before the pandemic period.

### 5.3 Study selection process

The searches conducted to retrieve pandemic-period evidence yielded a total of 5,549 records, one additional record was identified through personal communication, resulting in a total of 5,550 records. Records were imported into an Endnote database library and duplicates were removed. After deduplication, a total of 5,165 records remained. The title and abstract of the 5,165 records were screened by one reviewer and if relevant, the full text was also screened using the eligibility criteria from section 5.1. A second reviewer consistency checked all the studies selected for inclusion. If disagreements arose, these were discussed, and a third reviewer was consulted to make a final inclusion decision. In relation to the pandemic period, three records met the inclusion criteria for this rapid review (two qualitative papers and one ongoing trial of interventions).

For the pre-pandemic period literature, two independent reviewers screened title and abstracts. If relevant, the full text was reviewed by two independent reviewers for inclusion. If disagreements arose, these were discussed, and a third reviewer was consulted to make a final inclusion decision.

### 5.4 Data extraction

#### 5.4.1 Pandemic-related data extraction

One researcher performed the data extraction and a second researcher carried out consistency checks. The following information was extracted when reported:

- Citation
- Country
- Study design
- Data collection methods
- Sample size
- Population
- Setting
- Dates of data collection
- Key findings

A comments column was added to report key information that was not captured above and to record any limitations of the included primary sources.

#### 5.4.2 Pre-pandemic data extraction

One researcher performed the data extraction and a second researcher carried out consistency checks. The following information was extracted when reported:

- Citation
- Review period
- Review purpose
- Included study designs
- Quality rating
- Included outcome measures
- Number of included studies
- Key characteristics
- Findings and observations/notes

### 5.5 Quality appraisal

Quality assessment was undertaken by a single reviewer, with verification of all judgements by a second reviewer. Any discrepancies were discussed and resolved amongst the review team. Critical appraisal of pandemic period primary studies was conducted using the Critical Appraisal Skills Programme (CASP) tool for qualitative research (Critical Appraisal Skills Programme, 2018).

Critical appraisal of pre-pandemic systematic reviews was conducted using the AMSTAR 2 tool (Shea et al., 2017). The results of the quality appraisal for pre-pandemic studies can be seen in table 3. The reviewers agreed to give prominence to five critical domains on the AMSTAR 2 tool (see below) that could affect the validity of a review and its conclusions.

Critical domains:

- Protocol registered before commencement of the review (item 2)
- Adequacy of the literature search (item 4)
- Justification for excluding individual studies (item 7)
- Risk of bias from individual studies being included in the review (item 9)
- Consideration of risk of bias when interpreting the results of the review (item 13)

### 5.6 Synthesis

Barriers and facilitators to screening uptake identified relating to the COVID-19 pandemic period were compared against the barriers and facilitators identified before the pandemic. Where possible these were examined according to cancer type and population type. Data was synthesised narratively to provide a collective interpretation of the evidence.

## 6. EVIDENCE

### 6.1 Study selection flow chart for pandemic literature

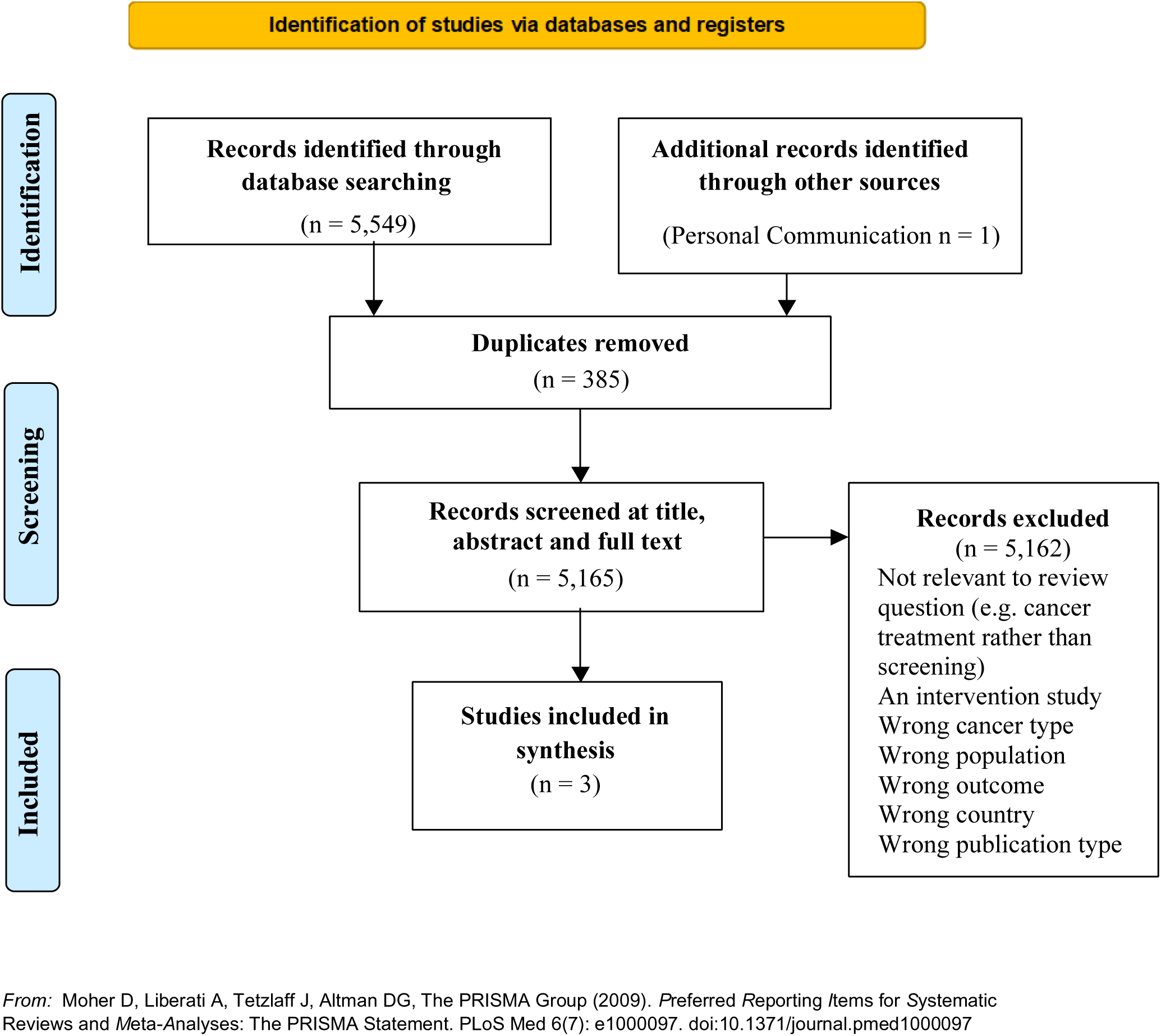

## 7.2 Acknowledgements

The authors would like to thank Hayley Heard, Sharon Hillier, Sunil Dolwani, Sikha de Souza, Rashmi Kumar and Stephanie Smits for their contributions during stakeholder meetings in guiding the focus of the review and interpretation of findings.

## 8. ABOUT THE WALES COVID-19 EVIDENCE CENTRE (WCEC)

The WCEC integrates with worldwide efforts to synthesise and mobilise knowledge from research.

We operate with a core team as part of Health and Care Research Wales, are hosted in the Wales Centre for Primary and Emergency Care Research (PRIME), and are led by Professor Adrian Edwards of Cardiff University.

The core team of the centre works closely with collaborating partners in Health Technology Wales, Wales Centre for Evidence-Based Care, Specialist Unit for Review

Evidence centre, SAIL Databank, Bangor Institute for Health & Medical Research/ Health and Care Economics Cymru, and the Public Health Wales Observatory.

Together we aim to provide around 50 reviews per year, answering the priority questions for policy and practice in Wales as we meet the demands of the pandemic and its impacts.

**Director:**

Professor Adrian Edwards

**Contact Email:**

**Website:**

https://healthandcareresearchwales.org/about-research-community/wales-covid-19-evidence-centre

## 9. APPENDIX

## APPENDIX 1: Resources searched during Rapid Review Searching

A single list of resources has been developed for guiding and documenting the sources searched as part of a Rapid Review. All ‘core’ resources should be searched, but other resources may be considered if appropriate to the topic, or time allows.

For those resources used, record the search strategies used below the table.

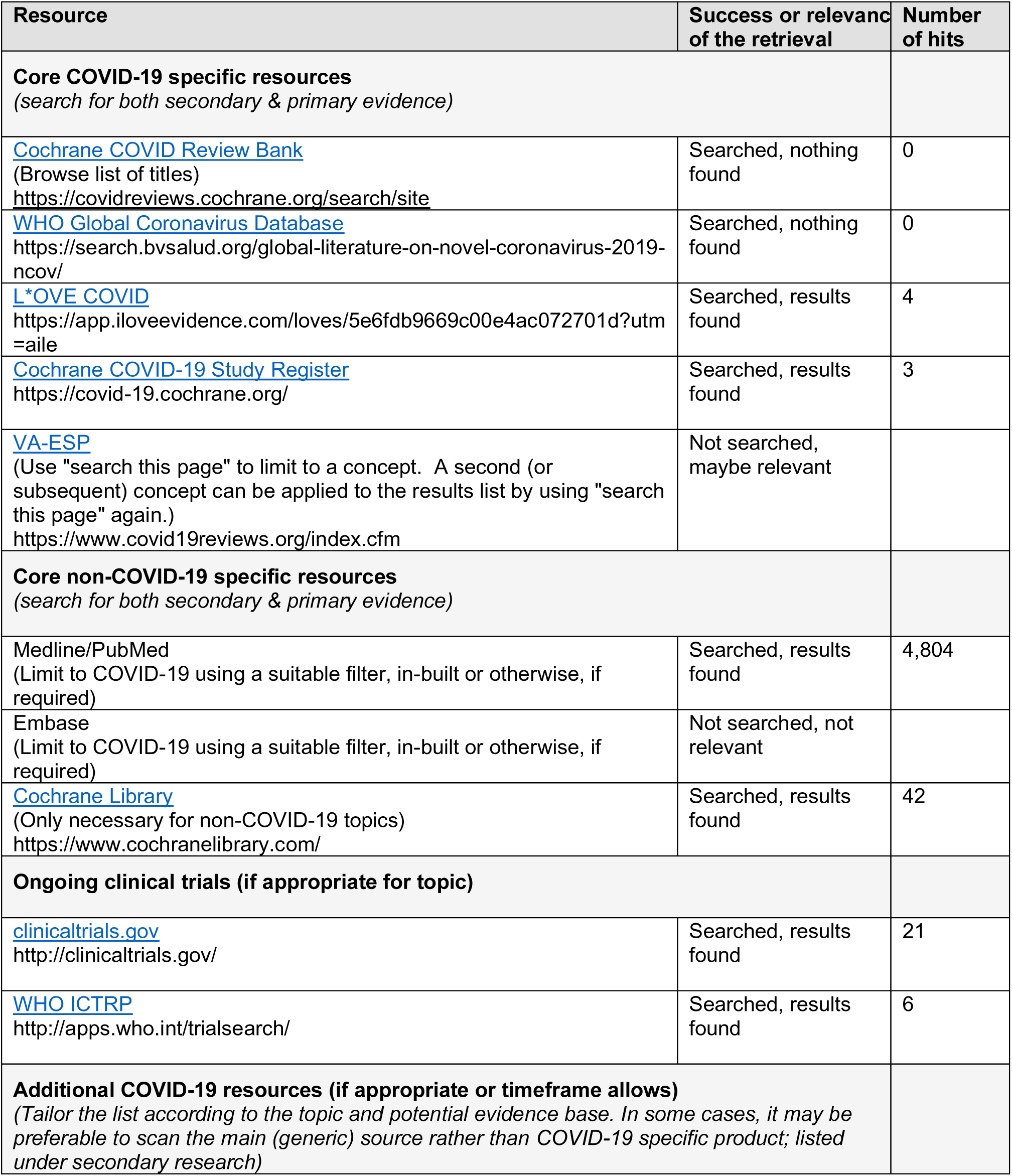
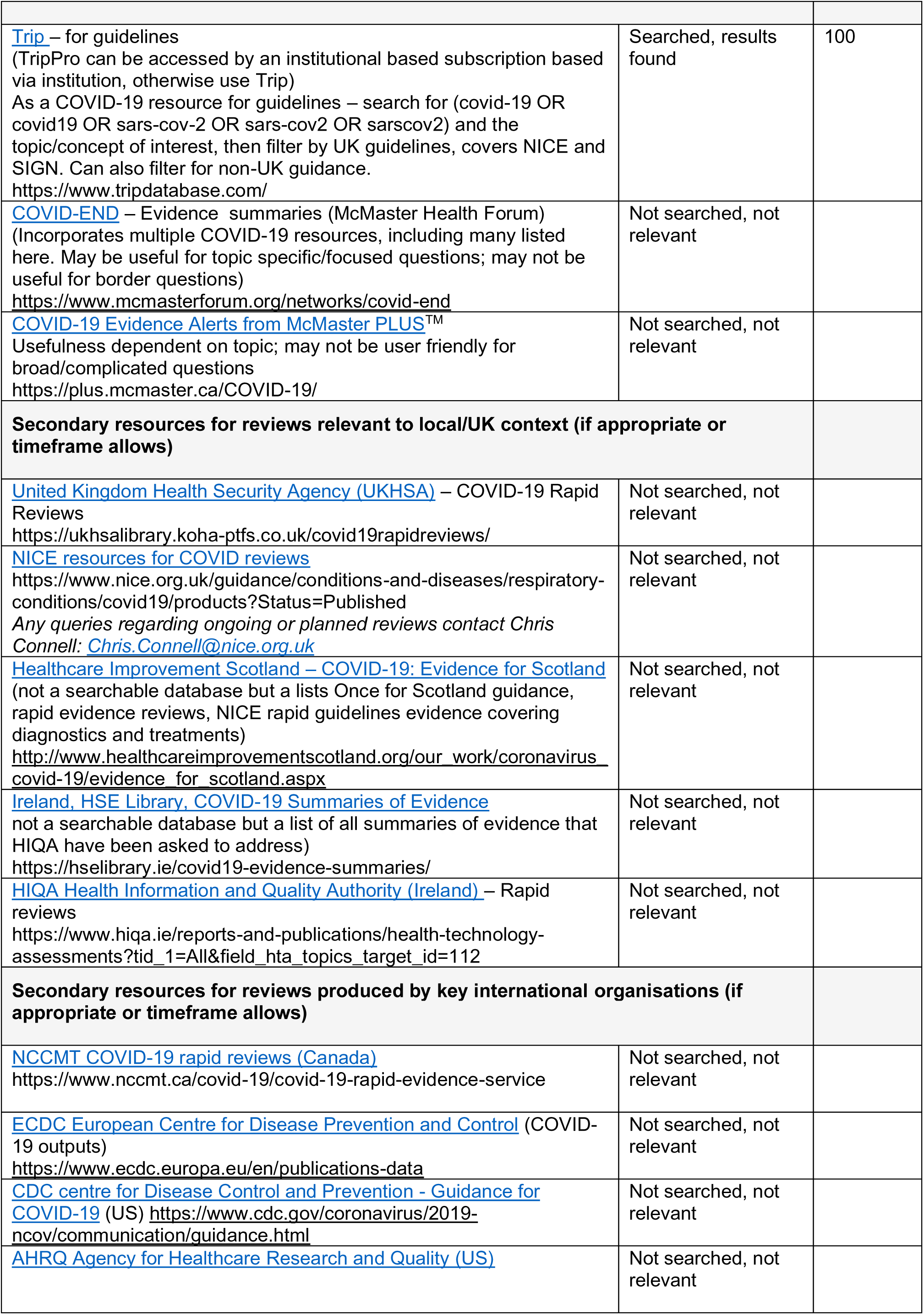
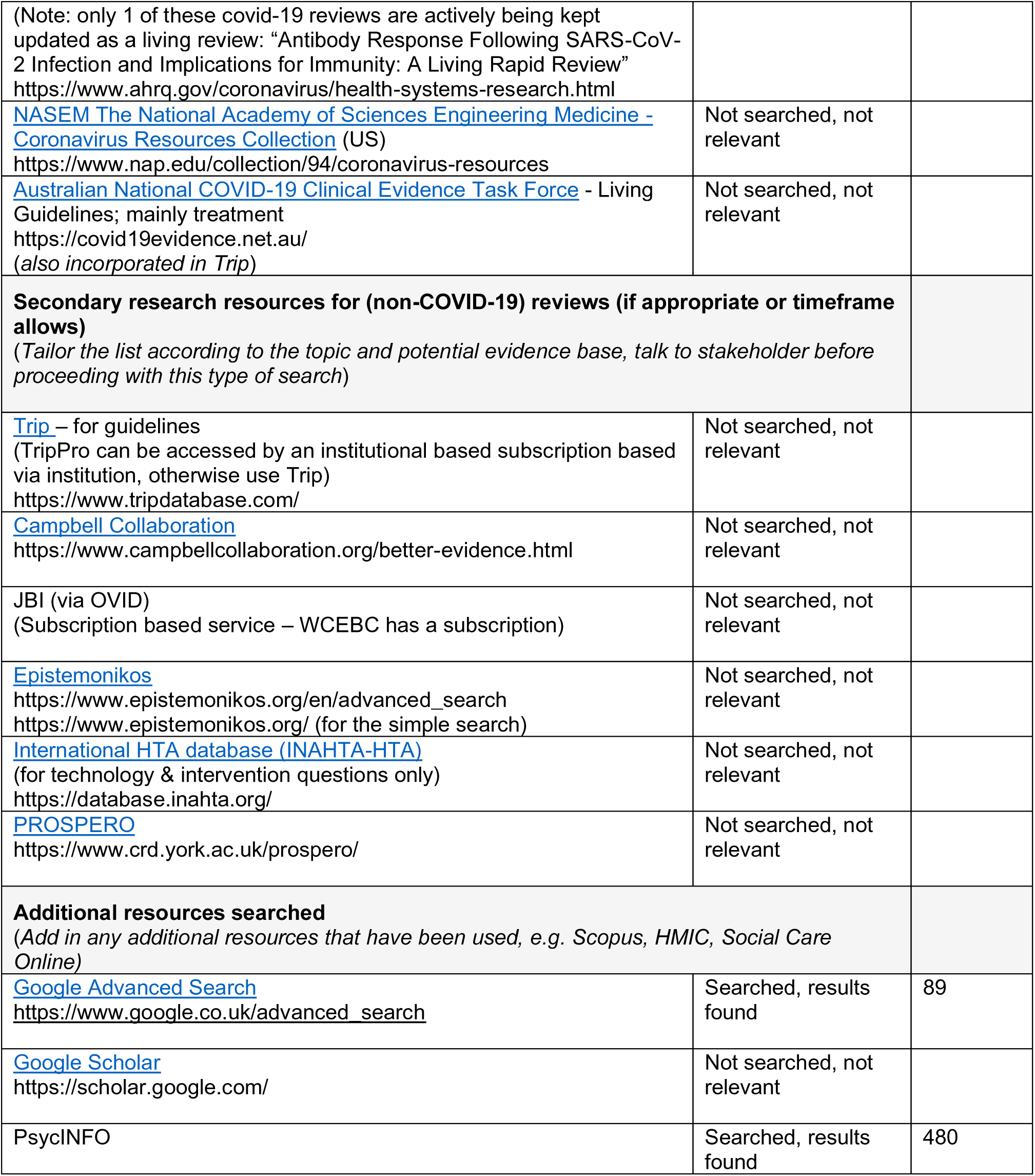

## Appendix 2. MEDLINE search strategy

| Set# | Searched for | Results |
| --- | --- | --- |
| S1 | (((((MESH.EXACT.EXPLODE("Early Detection of Cancer") OR MESH.EXACT.EXPLODE("Mass Screening"))))) | 165852 |
| S2 | (((((TI,ab(Screeni* OR "cancer screening" OR "early detection" OR "detection" OR "early diagnosis" OR "diagnosis" OR "cancer screening tests" OR "testing" OR "mass screening"))))) | 3493216 |
| S3 | (((((TI,ab(Mammogra* OR (colonoscop*) OR ("faecal immunochemical test") OR ("feecal immunochemical testing") OR (smear) OR (pap smear) OR (HPV) OR ("human papillomavirus"))))) | 202016 |
| S4 | S3 OR S2 OR S1 | 3634559 |
| S5 | (((((MESH.EXACT.EXPLODE("Breast Neoplasms") OR MESH.EXACT.EXPLODE("Neoplasms") OR MESH.EXACT("Uterine Cervical Neoplasms") OR MESH.EXACT.EXPLODE("Breast Cancer Lymphedema") OR MESH.EXACT.EXPLODE("Colorectal Neoplasms"))))) | 3692423 |
| S6 | (((((ti,ab("breast cancer" OR "bowel cancer" OR "cervical cancer" OR "colorectal cancer" OR "cancer" OR "neoplasm"))))) | 1997371 |
| S7 | S6 OR S5 | 4217020 |
| S8 | ((((((MESH.EXACT.EXPLODE("Minority Groups") OR MESH.EXACT.EXPLODE("Health Disparity, Minority and Vulnerable Populations") OR MESH.EXACT.EXPLODE("Minority Health") OR MESH.EXACT.EXPLODE("Vulnerable Populations") OR MESH.EXACT.EXPLODE("Sexual and Gender Minorities") OR MESH.EXACT.EXPLODE("Ethnic and Racial Minorities") OR MESH.EXACT.EXPLODE("Risk Factors") OR MESH.EXACT.EXPLODE("Poverty Areas") OR MESH.EXACT.EXPLODE("Poverty") OR MESH.EXACT.EXPLODE("Disabled Persons") OR MESH.EXACT.EXPLODE("Age Factors")))))) | 1071724 |
| S9 | (((((TI,ab(minorit* OR vulnerable OR ethnic* OR Black OR Asian OR LGBT* OR homosexual* OR bisexual* OR race OR gender OR ("hard to reach") OR underserved OR age OR ("travelling communit*") OR ("disabled person*") OR disab* OR poverty OR ("working poor") OR emigrant* OR immigrant* OR refugee OR ("asylum seeker") OR gypsy OR Roma OR ("Irish traveller"))))) | 3488651 |
| S10 | ((((((MESH.EXACT.EXPLODE("Socioeconomic Factors") OR MESH.EXACT.EXPLODE("Health Status Disparities") OR MESH.EXACT.EXPLODE("Healthcare Disparities") OR MESH.EXACT.EXPLODE("Medically Underserved Area") OR MESH.EXACT.EXPLODE("Gender Equity") OR MESH.EXACT.EXPLODE("Sociological Factors") OR MESH.EXACT("Age Factors") OR MESH.EXACT("Sex Factors") OR MESH.EXACT.EXPLODE("Health Equity")))))) | 1572980 |
| S11 | (((((Ti,ab(("social disparit*") OR ("social inequ*") OR ("health disparit*") OR ("health inequ*") OR ("healthcare disparit*") OR ("health status disparit*") OR (disparit*) OR (inequ*) OR (equali*) OR ("disparities in health") OR ("socioeconomically disadvantaged") OR ("economic level") OR ("social class") OR ("social determinant") OR ("social status") OR ("social background") OR ("social circumstance") OR ("socio-economic") OR (socioeconomic) OR (sociodemographic) OR (socio-demographic)))))))))) | 355366 |
| S12 | S11 OR S10 OR S9 OR S8 | 5076729 |
| S13 | (((((Ti,ab (compliance) OR (comply*) OR (adher*) OR (encourage*) OR (promot*) OR (uptake) OR (non-attend*) OR (nonattend*) OR (accept*) OR (attend*) OR (attitude*) OR (utilisation) OR (utilization) OR (refus*) OR (reluctan*) OR (non-respon*) OR (nonrespon*) OR (barriers) OR (facilitators) OR (enabl*) OR (engag*) OR (perception*) OR (experience*) OR (views) OR (intention*))))) | 5881849 |
| S14 | ((((interview* OR ("focus group*") OR discussion OR survey* OR observation*))) | 2915954 |
| S15 | ((((Qualitative OR ("mixed method*") OR cohort))) | 1176329 |
| S16 | S15 OR S14 | 3733195 |
| S17 | (S16 AND S13 AND S12 AND S7 AND S4) and (pd(2020-2029)) | 4804° |

## Appendix 3. CASP Checklists

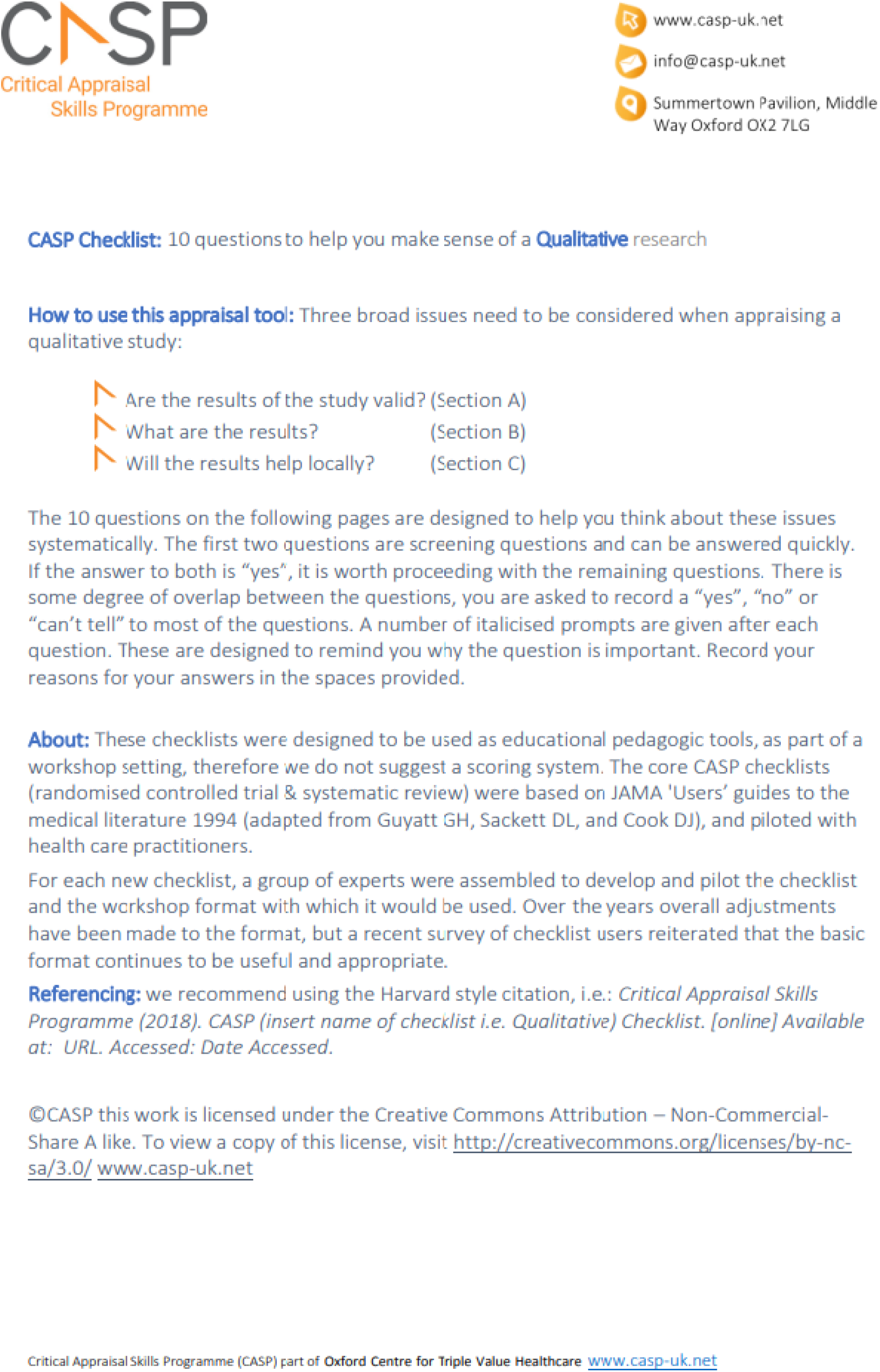
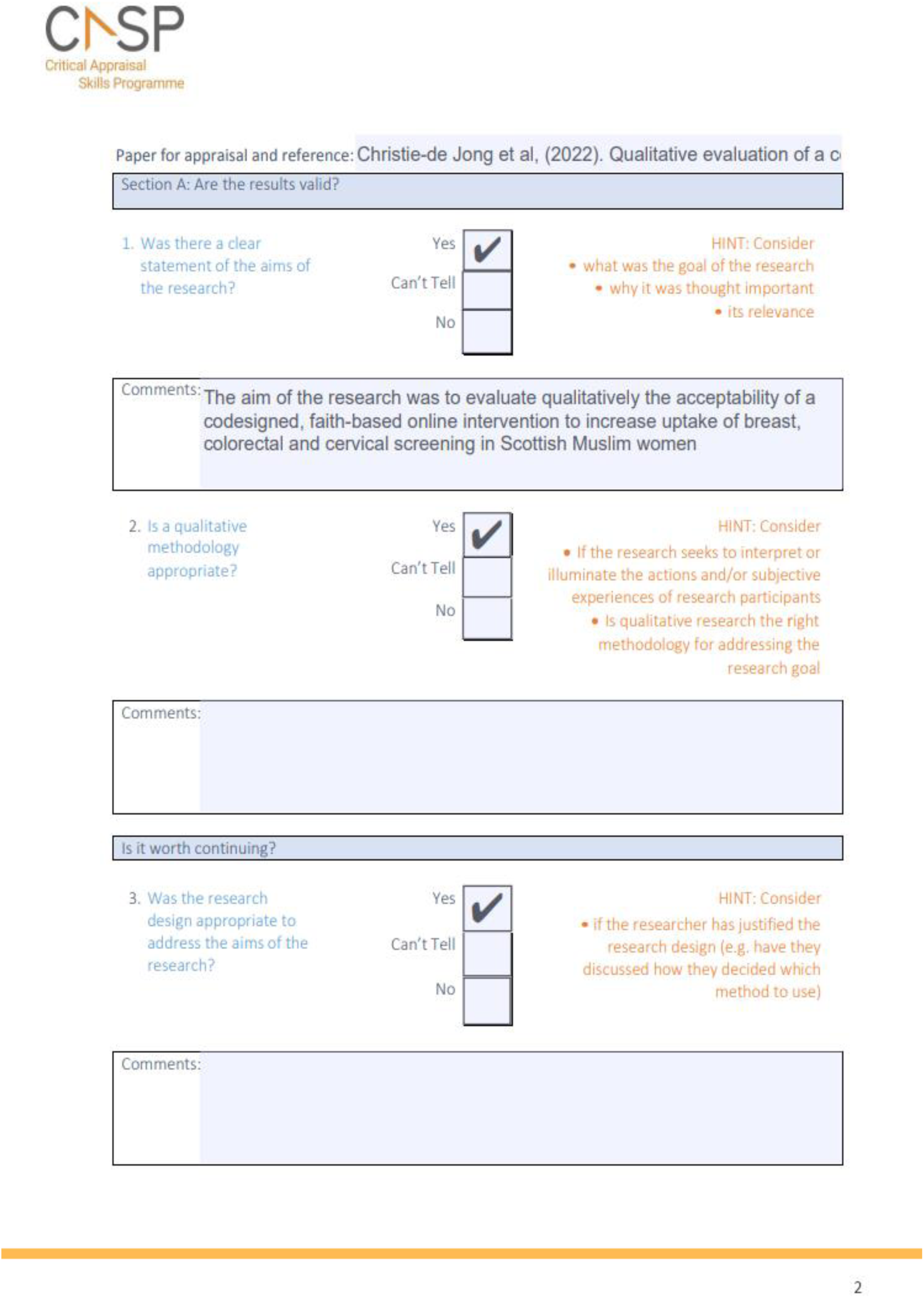
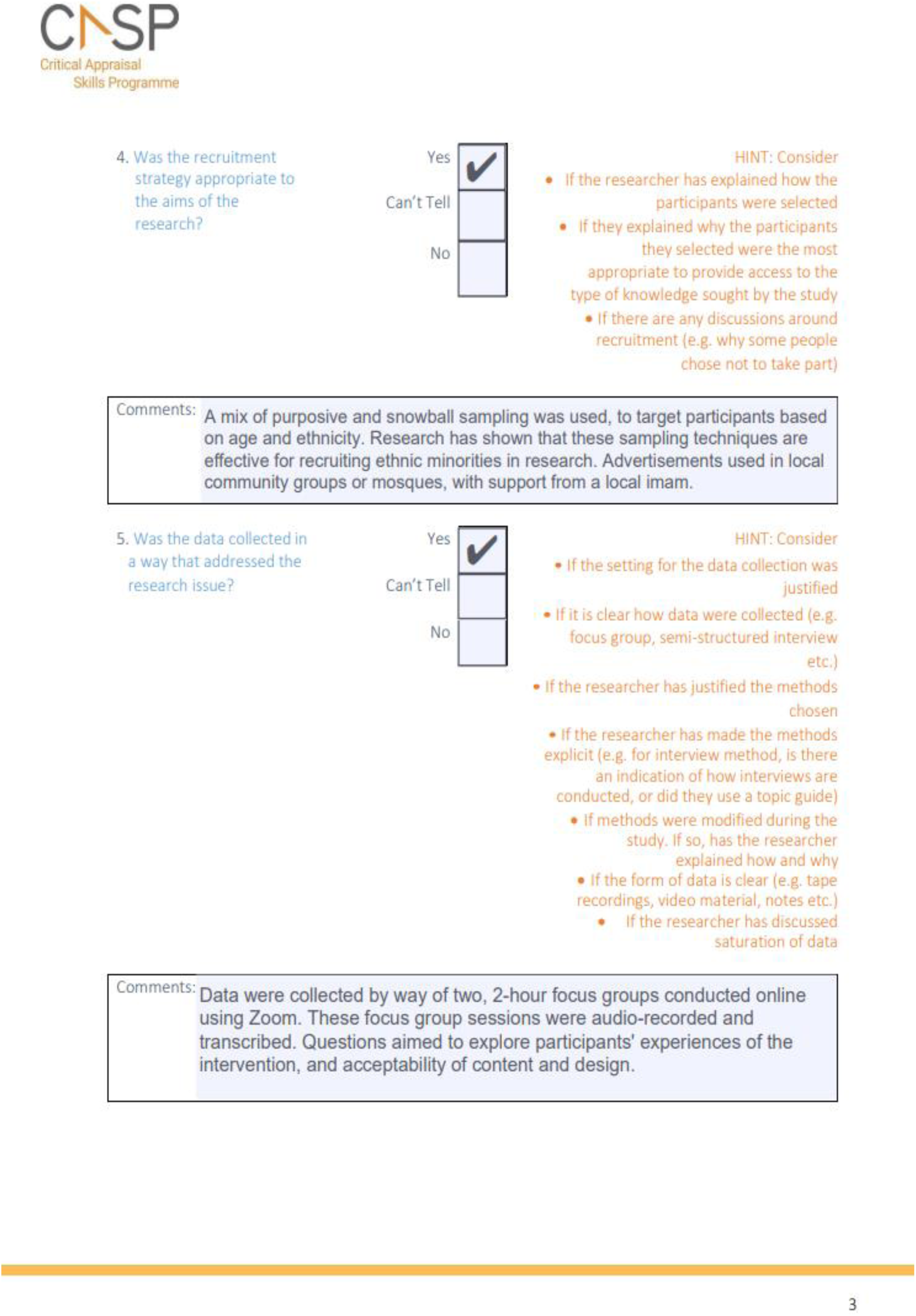
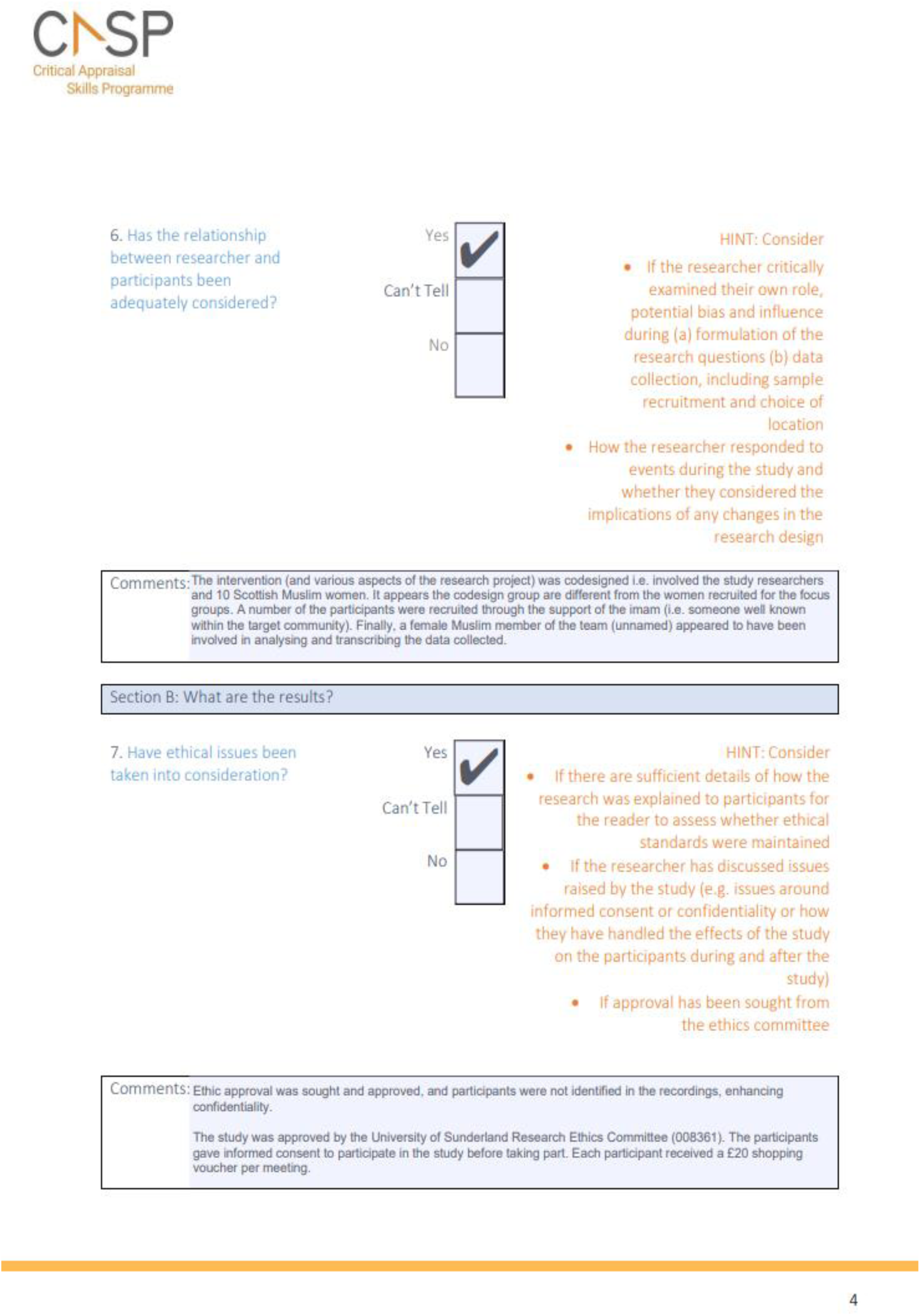
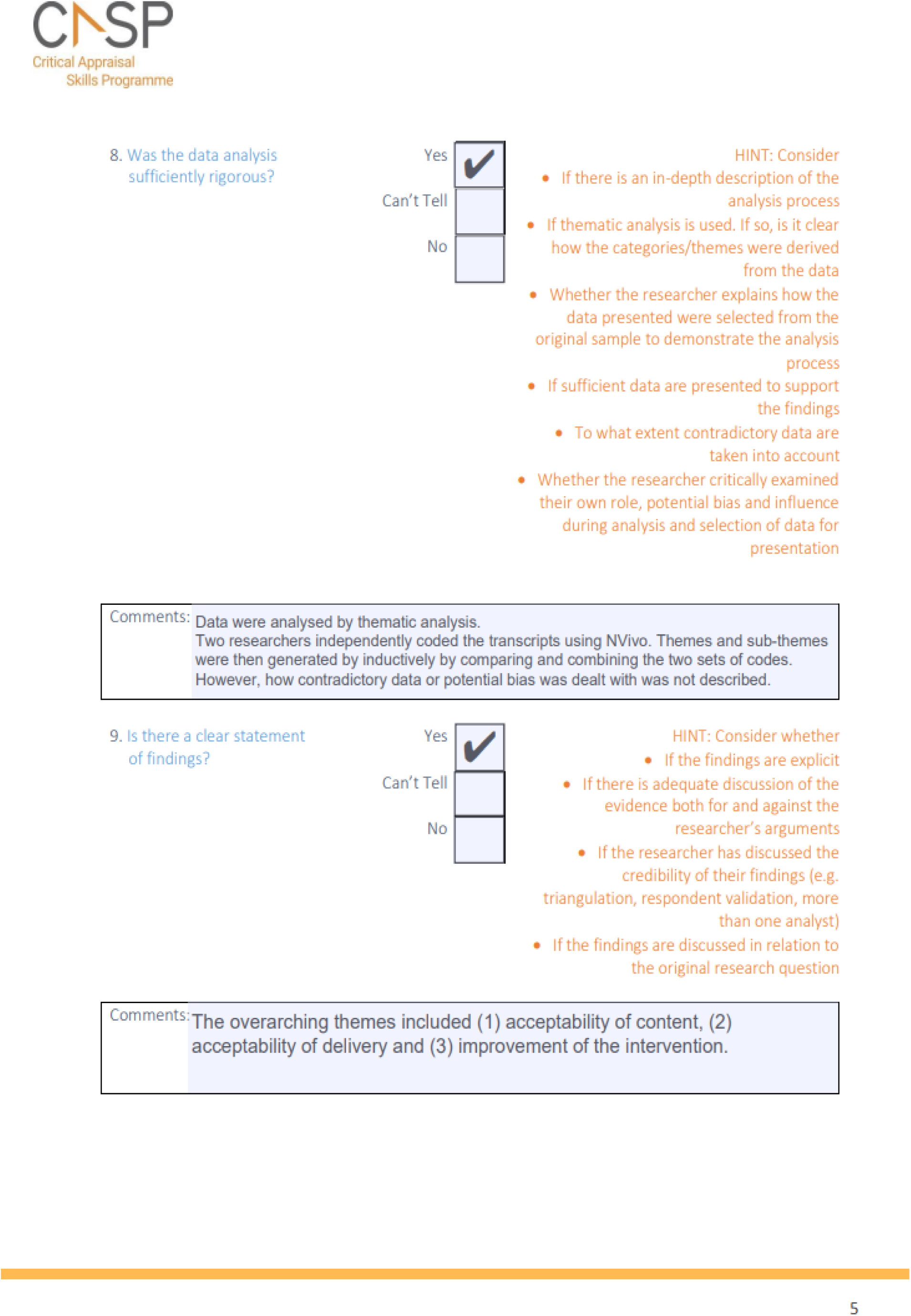
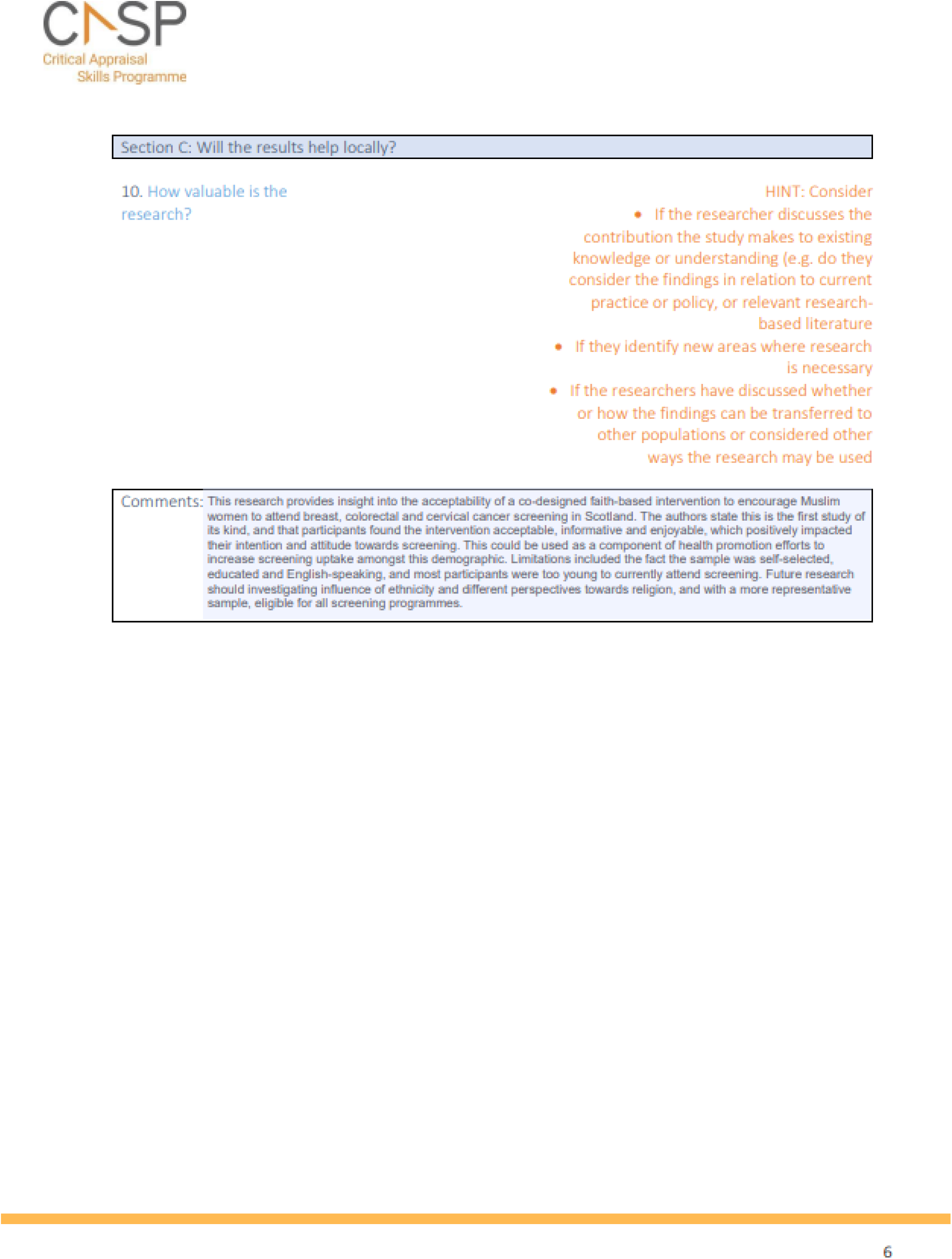
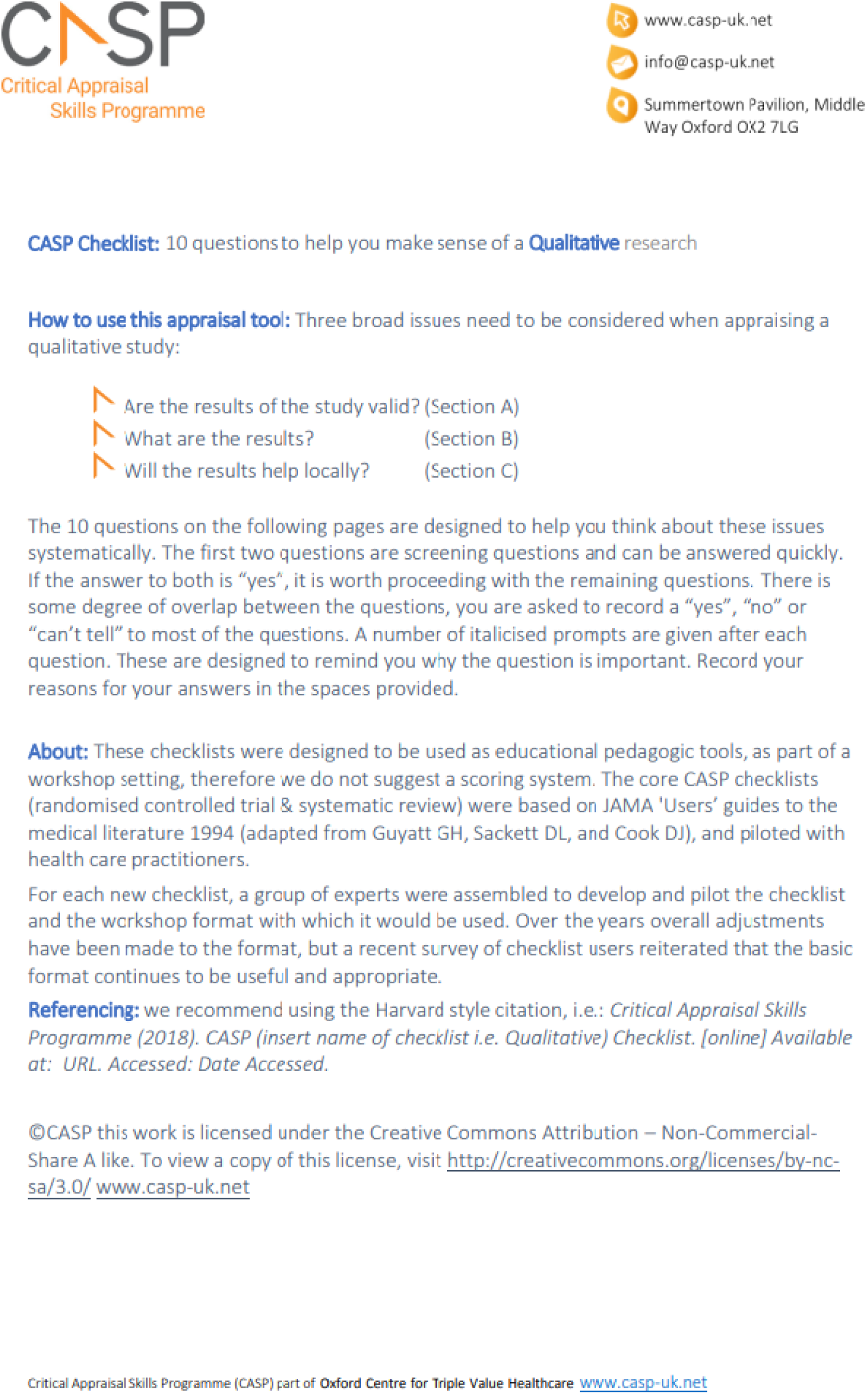
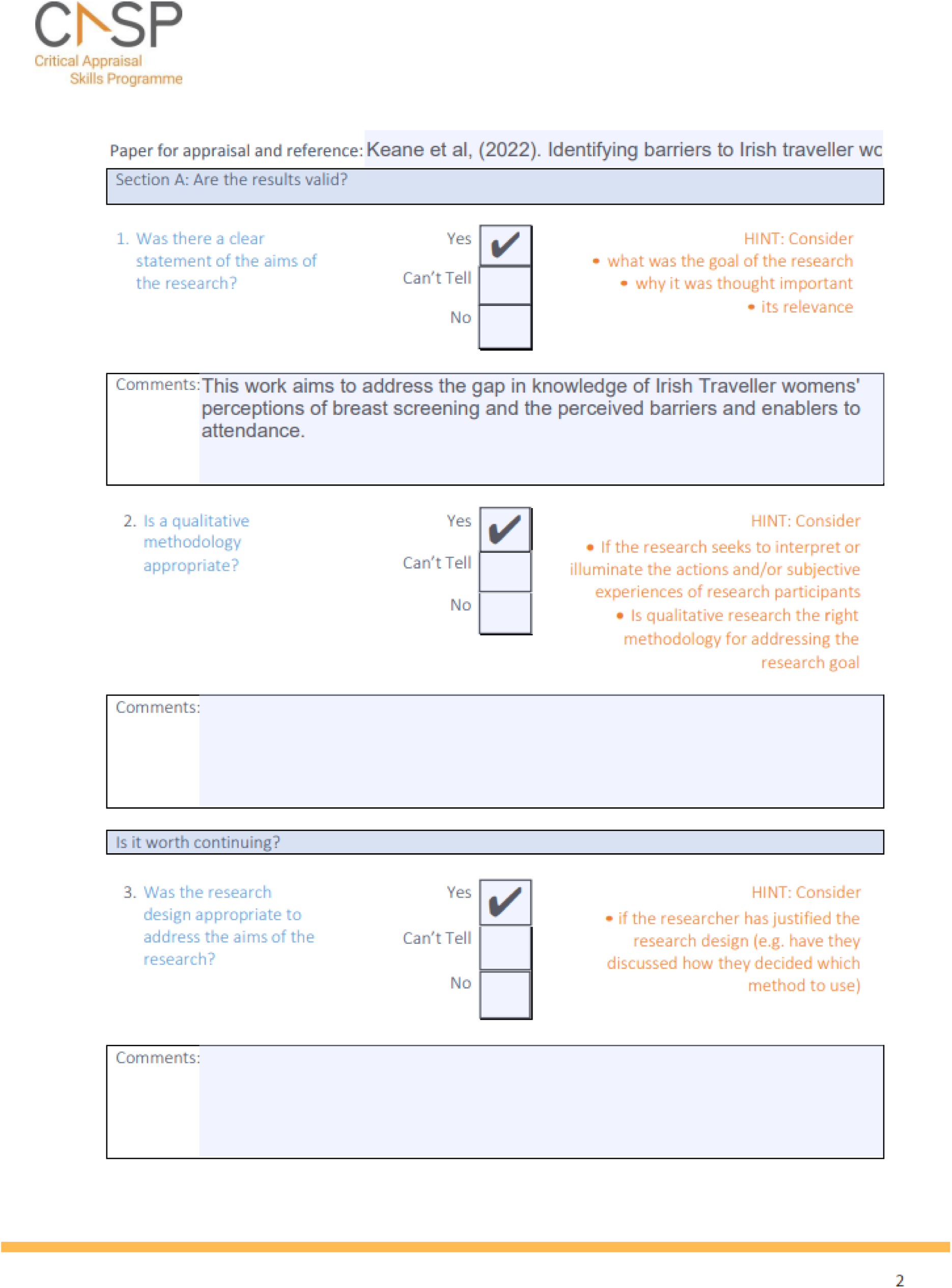
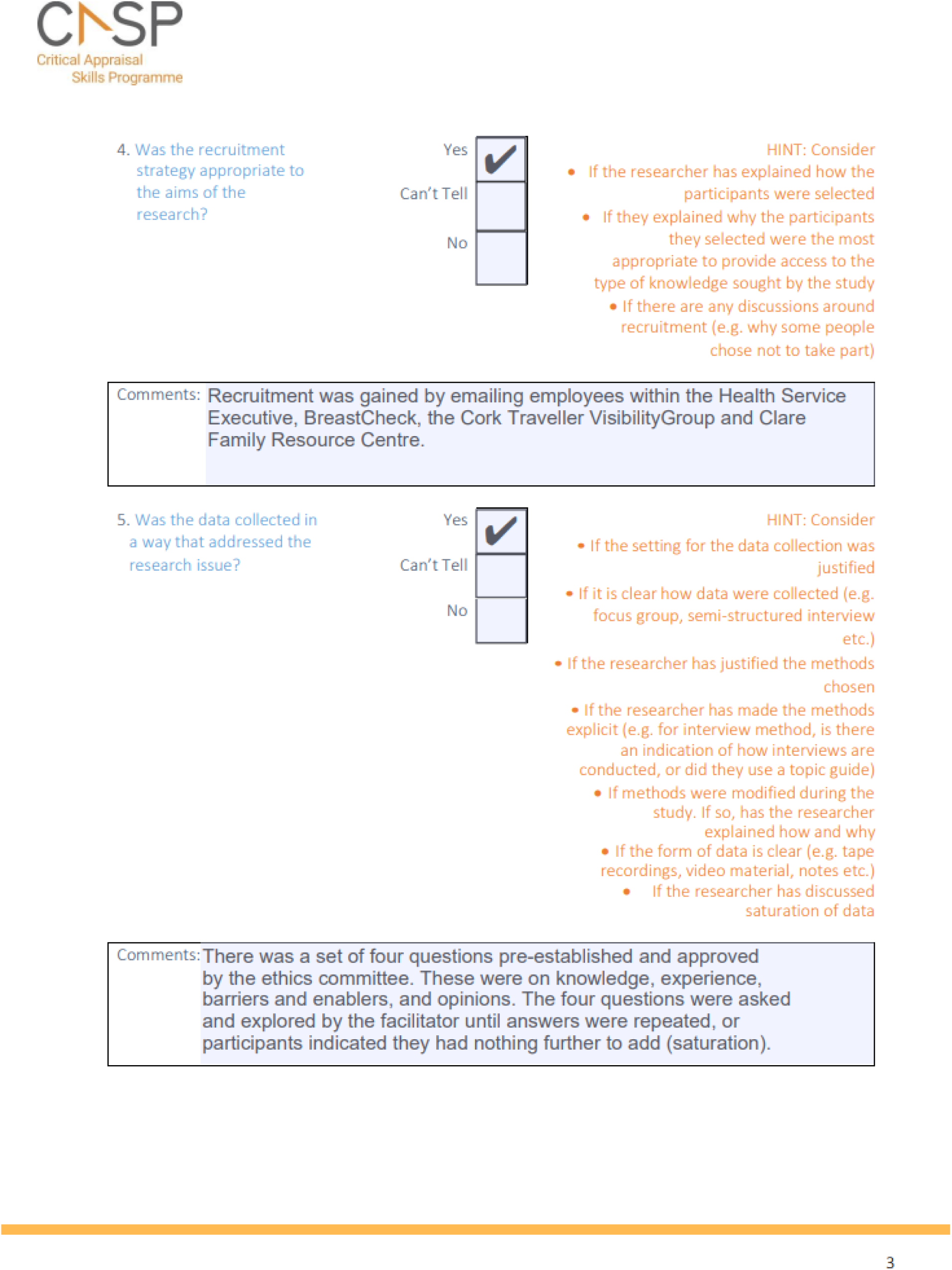
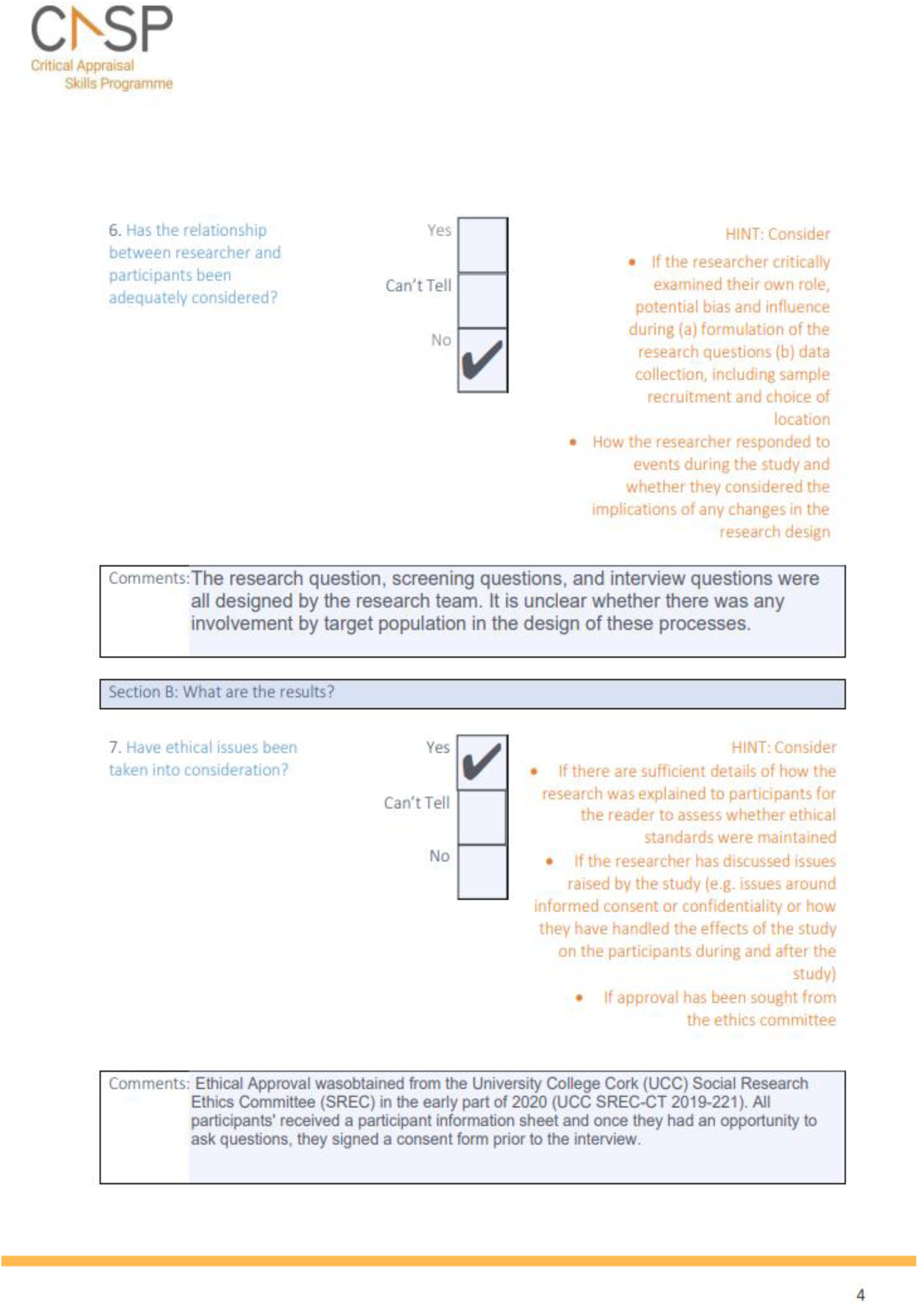
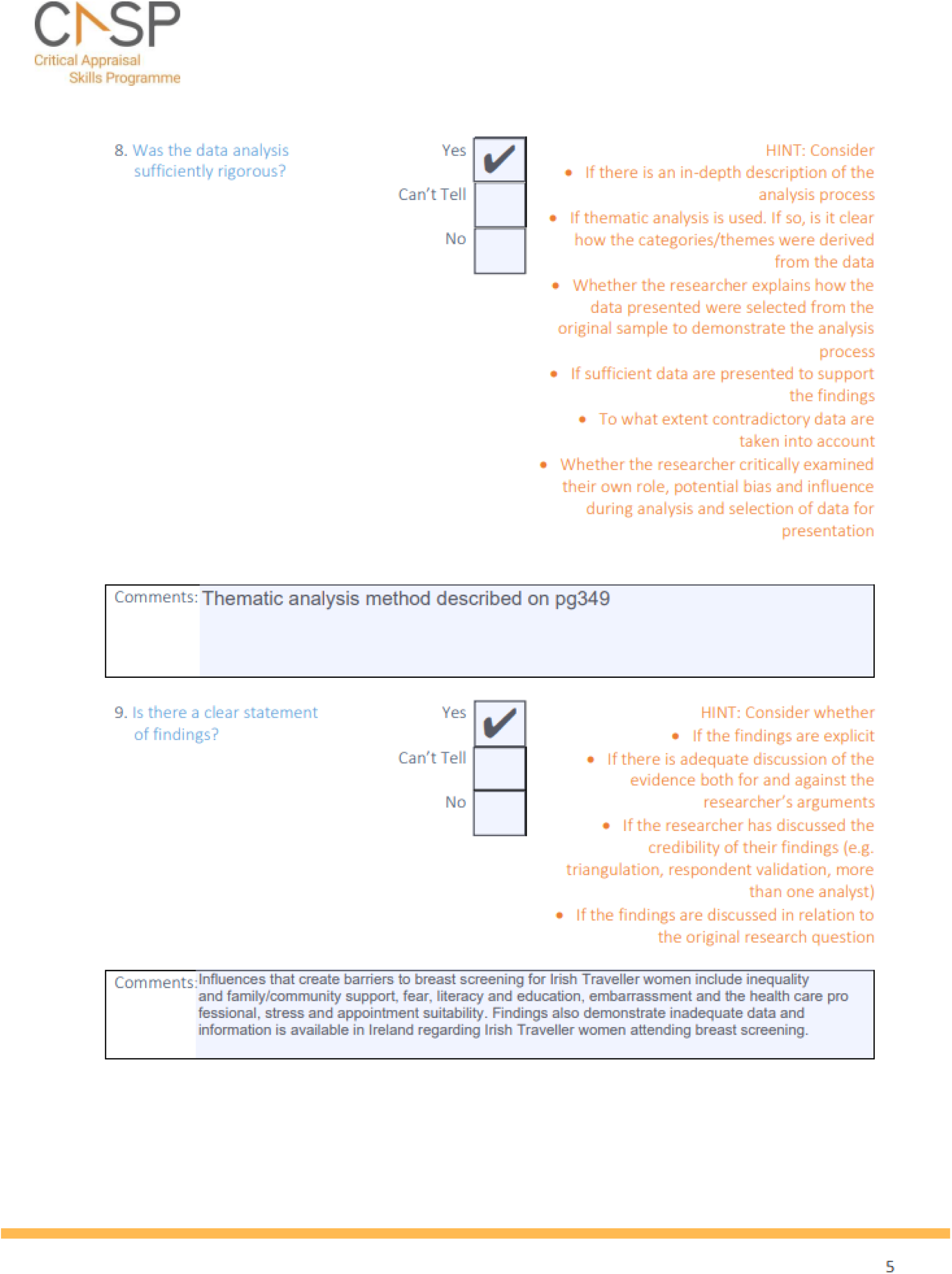
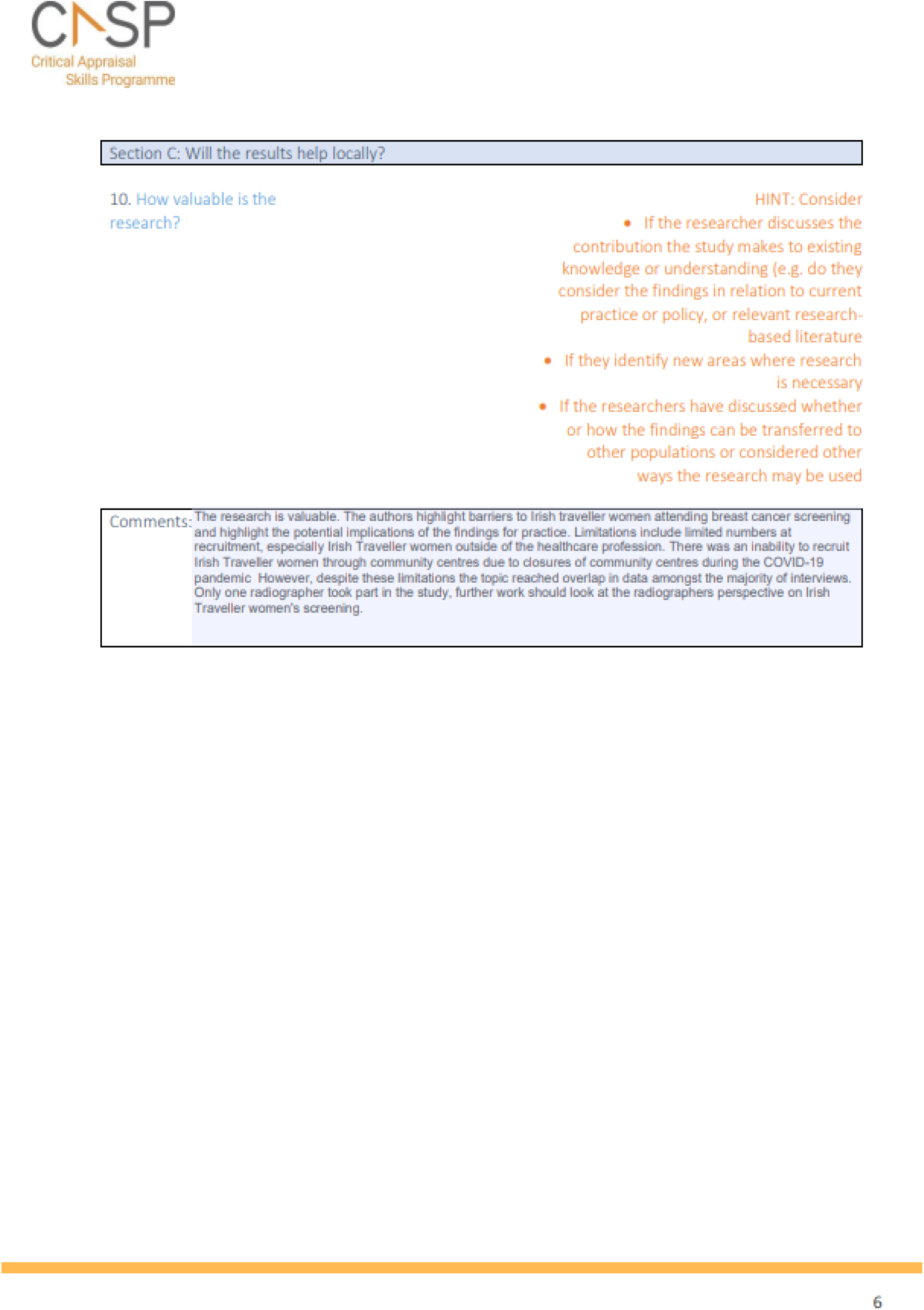

## Abbreviations

Acronym: Full Description
AMSTAR: A Measurement Tool to Assess Systematic Reviews
BAME: Black, Asian and Minority Ethnic
CASP: Critical Appraisal Skills Programme
CCGs: Clinical Commissioning Groups
COVID-19: Coronavirus disease 2019
CRC: Colorectal Cancer
FSS: Flexible Sigmoidoscopy Screening
GPs: General Practitioners
LGBT: Lesbian, Gay, Bisexual and Trans
NHS: National Health Service
RCT: Randomised Control Trial
RES: Rapid Evidence Summary
SES: Socioeconomic Status
UK: United Kingdom
USA: United States of America
WCEC: Welsh Covid-19 Evidence Centre
WHO: World Health Organisation

